# European Fitness Landscape in Children and Adolescents: updated reference values, fitness maps, and country rankings based on nearly 8 million data points from 34 countries gathered by the FitBack network

**DOI:** 10.1101/2022.06.09.22275139

**Authors:** Francisco B Ortega, Bojan Leskošek, Rok Blagus, Jose J. Gil-Cosano, Jarek Mäestu, Grant Tomkinson, Jonatan R. Ruiz, Evelin Mäestu, Gregor Starc, Ivana Milanovic, Tuija H. Tammelin, Maroje Sorić, Claude Scheuer, Attilio Carraro, Mónika Kaj, Tamás Csányi, Luis B. Sardinha, Matthieu Lenoir, Arunas Emeljanovas, Brigita Mieziene, Labros S. Sidossis, Maret Pihu, Nicola Lovecchio, Kenn Konstabel, Konstantinos D. Tambalis, Lovro Štefan, Clemens Drenowatz, Lukáš Rubín, Seryozha Gontarev, José Castro-Piñero, Jérémy Vanhelst, Brendan O’Keeffe, Oscar L. Veiga, Thordis Gisladottir, Gavin Sandercock, Marjeta Misigoj-Durakovic, Claudia Niessner, Eva-Maria Riso, Stevo Popovic, Saima Kuu, Mai Chinapaw, Iván Clavel, Idoia Labayen, Janusz Dobosz, Dario Colella, Susi Kriemler, Sanja Salaj, Maria Jose Noriega, Klaus Bös, Mairena Sánchez-López, Timo A. Lakka, Garden Tabacchi, Dario Novak, Wolfgang Ahrens, Niels Wedderkopp, Gregor Jurak, the FitBack, HELENA and IDEFICS consortia

## Abstract

**Objectives:** (1) To develop reference values for health-related fitness European children and adolescents aged 6–18 years that are the foundation for the web-based, open-access and multi-language fitness platform (FitBack); (2) To provide comparisons across European countries.

**Methods:** This study builds on a previous large fitness reference study in European youth by: (1) widening the age demographic, (2) identifying the most recent and representative country-level data, and (3) including national data from existing fitness surveillance and monitoring systems. We used the ALPHA test battery as it comprises tests with the highest test-retest reliability, criterion/construct validity, and health-related predictive validity: the 20-m shuttle run (cardiorespiratory fitness); handgrip strength and standing long jump (muscular strength); and body height, body mass, body mass index, and waist circumference (anthropometry). Percentile values were obtained using the GAMLSS method.

**Results:** A total of 7,966,693 data points from 34 countries (106 datasets) were used to develop sex- and age-specific percentile values. In addition, country-level rankings based on mean percentiles are provided for each fitness test, as well as an overall fitness ranking. Finally, an interactive fitness platform, including individual and group reporting, and European fitness maps, is provided and freely available at www.fitbackeurope.eu.

**Conclusions:** This study discusses the major implications of fitness assessment in youth from a health, educational and sport perspective, and how the FitBack reference values and interactive web-based platform contribute to it. Fitness testing can be conducted in school and/or sport settings, and the interpreted results be integrated in the healthcare systems across Europe.

**What is already known on this topic:**

- Fitness testing in youth is important from a health, educational and sport point of view.
- The EU-funded ALPHA project reviewed the existing evidence and proposed a selection of field-based fitness tests that showed the highest test-retest reliability, criterion/construct validity, and health-related predictive validity among available tests.

**What this study adds:**

- The FitBack project provides the most up-to-date and geographically diverse reference fitness values for 6-to 18-year-old Europeans.
- This study introduces the first web-based, open-access, and multi-lingual fitness reporting platform (FitBack) providing interactive information and visual mapping of the European fitness landscape.

**How this study might affect research, practice, or policy:**

- From a health perspective, very low fitness levels are a non-invasive indicator of poor health at both the individual and group level (e.g., school, region), which have utility for health screening and may guide public health policy. There are already examples of regional and national fitness testing systems that are integrated into the healthcare systems.
- From an educational perspective, fitness testing is part of the school curriculum in many countries, and the FitBack platform offers physical education teachers an easy-to-use tool for interpreting fitness test results by sex and age.
- From a sport perspective, these reference values can help identify young individuals who are talented in specific fitness components.

## INTRODUCTION

Robust and consistent evidence supports that physical fitness is a powerful marker of health in children and adolescents [1,2]. Among the different fitness components, cardiorespiratory fitness (CRF, used in the literature and this article interchangeably with aerobic fitness) and muscular strength (used in the literature and this article interchangeably with muscular fitness) have shown the strongest and most consistent health-related associations, and are therefore considered to be health-related [3,4]. Other fitness components include muscular endurance, flexibility, motor fitness, and body composition/anthropometry (height, body mass, body mass index (BMI), and waist circumference). Recently, data from large registries have added compelling evidence linking both CRF and muscular strength in late adolescence with all-cause mortality and cardiovascular- and cancer-specific mortality in later life [5–8]. In addition, these two fitness components predict severe, chronic, and irreversible all-cause disease 30 years later as indicated by granted disability pensions [9–12], and also specifically cardiovascular, musculoskeletal, neurological, and psychiatric diseases granted by a disability pension [9–12]. Particularly, CRF is the most well-studied and strongest predictor of future health. Indeed, a position stand from the American Heart Association has highlighted the clinical value of CRF in youth and recommended that it be regularly assessed [13].

In addition to the well-documented associations between fitness and physical/mental health among youth [1–4,14], emerging evidence supports that better fitness is related to better cognition, academic performance, and healthier structural and functional brain outcomes [15–29]. For example, recent observations from the ActiveBrains project have shown that whole brain size, as well as total gray and white matter volumes, is larger in fit compared to unfit children with overweight/obesity [30]. This is important because brain size is positively associated with intelligence [31].

This evidence begs the question: what are the best methods to assess health-related fitness among children and adolescents? The EU-funded ALPHA project was designed to answer this question. By conducting a set of systematic reviews [2,32,33] and methodological papers, the ALPHA consortium aimed to identify which field-based fitness tests demonstrated the highest test-retest reliability, criterion/construct validity, and health-related predictive validity (see ALPHA summary article [34]). Anthropometry and body composition were tightly linked to fitness performance and health, and were therefore considered as fitness components in the ALPHA project. The final output of the project was the ALPHA-fitness test battery for children and adolescents, which in its High-Priority version (a shorter, more suitable version for school-based use) recommended using: the 20-m shuttle run test for assessing CRF; the handgrip and standing long jump tests for assessing muscular strength and power; and BMI and waist circumference as indicators of total and central obesity. A year later and after following a similar systematic review process, the U.S. Institute of Medicine (now the National Academy of Medicine) recommended these tests for the assessment of youth physical fitness [35,36], strengthening the recommendation of using these selected tests.

The EU-funded the FitBack consortium (www.fitbackeurope.eu) titled the European Network for the Support of Development of Systems for Monitoring Physical Fitness of Children and Adolescents. The major goal of the network is to take an important step toward the implementation of fitness surveillance and monitoring across Europe as an educational tool for physical literacy[37]. Physical literacy can be defined as ‘the motivation, confidence, physical competence, knowledge and understanding to value, and take responsibility for, maintaining purposeful physical pursuits/activities throughout the life-course”[38]. In this context, fitness testing should be much more than just ‘one more school assessment’. Schools are in a unique position to positively affect the physical activity and physical fitness levels of their students not only in the short term, but also by instilling values and skills that will help children throughout their lives.

The final output of the FitBack project has been the development of a web-based, open-access, and multi-language fitness platform, which allows the results of fitness testing to be automatically and interactively interpreted based on sex- and age-specific reference values, and is supported by user-friendly visual feedback and tips for improvement. For this purpose, we gathered available fitness data on European children and adolescents, accumulating 8 million data points to create reference values for European children and adolescents aged 6 to 18 years.

The aim of this article is to present the most comprehensive and up-to-date health-related fitness reference values for European children and adolescents. Additionally, we provide European fitness maps for the main health-related fitness components. Since pediatric obesity is being comprehensively monitored by other organizations (e.g., World Obesity Federation www.worldobesity.org/, WHO-Europe www.euro.who.int/en/health-topics/disease-prevention/nutrition/activities/who-european-childhood-obesity-surveillance-initiative-cosi), the focus of this article is mainly on CRF and muscular strength. Nonetheless, we also provide reference values and European maps for anthropometric measures (body height, body mass, BMI, and waist circumference) as online supplementary material.

## METHODS

### Data search and pooling

A systematic review of existing data sets including fitness tests in children and adolescents was previously performed by Tomkinson et al. and details of the search have been published [39]. These data were included in the FitBack dataset, with Monte Carlo simulation used to produce pseudodata (from reported means and SDs) when raw data were unavailable. In addition to this, the authors of the FitBack network conducted a narrative search based on fitness terms to identify new datasets not included in the Tomkinson et al. review [39]. For inclusion, valid data on sex, age and at least one of the ALPHA fitness tests (High-Priority version) was required. In the previous study by Tomkinson et al., the age range was 9-to 17-year-olds, whereas in this study we widened the age demographic to include 6-to 18-year-olds. It is important to note that our search strategy was fitness focused, and specific searches on adiposity, BMI, or waist circumference were not conducted for pragmatic reasons (e.g., the very large number of studies including these key words). Therefore, it is possible that we missed relevant anthropometry-specific datasets. This, together with the fact that other organizations are comprehensively monitoring pediatric obesity, is the reason why we primarily focused on CRF and muscular strength, and reported results for anthropometric measures (body height, body mass, BMI, and waist circumference) as online supplementary material.

The FitBack network involved many experienced researchers working in pediatric fitness across Europe, which helped to identify unpublished fitness datasets that were pooled with the gathered data. Moreover, massive data from existing surveillance systems in Europe were also included. Further, we excluded older datasets if a more recent and more representative dataset was available for certain countries. The ambition was to use the most recent available data for each country, which in some cases was a single large dataset, while in others was the accumulation of several studies or datasets covering different geographical regions within a country. Sources used for generating the reference values are available on the FitBack website (www.fitbackeurope.eu/en-us/fitness-map/sources) as well as in **Online Supplementary Table 1**. The entire Fitback procedures for pooling together existing fitness data were evaluated and approved by the Ethics Committee in Sports Science at the University of Ljubljana, Slovenia (the University coordinator the FitBack project).

### Physical fitness measures

The FitBack dataset was compiled for studies that used the ALPHA fitness test battery [2,32–34], since these tests have shown to be feasible, reliable, valid, and scalable for children and adolescents. Moreover, some of them are used in well-established European national fitness surveillance and monitoring systems, like SLOfit [40], NETFIT [41], and Fitescoula [42]. Specifically, CRF was assessed using the 20-m shuttle run test [43]. The number of completed stages was used as an indicator of CRF. However, different studies had expressed the result of the 20-m shuttle run test in other units, such as completed laps (shuttles) or speed at the last completed stage, and there are at least three known protocols/versions of this test [44]. All data were converted and harmonized into completed stages according to the original Léger’s protocol [43], as described elsewhere [44]. Muscular strength was assessed by the handgrip strength (i.e., upper-limb muscular strength) and standing long jump tests (i.e., lower-limb muscular strength). Total and abdominal adiposity were assessed by BMI and waist circumference, respectively, following standardized procedures. For handgrip, most studies collected data from both hands, with the average of the maxima for both hands used in our analyses. Two studies had handgrip strength data only for the dominant hand, which is known to be systematically higher compared to the non-dominant hand. Exploratory analyses on Spanish data in children [45] showed a 0.6 kg mean difference between hands and thus, we applied a −0.3 kg correction factor to these two studies to estimate the average score.

### Statistical analysis

We applied different cleansing procedures to the data. First, data were trimmed to remove values outside the probable lower and upper limits. The limits were defined based on authors’ experiences working with previous large datasets. The limits used were: 20-m shuttle run (0–21 stages), handgrip strength (0–80 kg), standing long jump (15–330 cm), body height (80–220 cm), body mass (0–200 kg), BMI (7–60 kg/m^2^), and waist circumference (40–130 cm). Second, outliers were identified and removed as follows. For each fitness measure, herein referred to as the *test*, a multivariate regression model including the *test* as the dependent variable and age (modelled as a cubic spline with 5 degrees of freedom), sex, and their interaction as independent variables was fitted. Studentized residuals were obtained and then 0.01% subjects with the smallest and largest studentized residuals were removed from further analysis. Weights were computed via iterative post-stratification (aka iterative proportional fitting) [46] to match the sample joint distributions by age, sex, and country to population data. Country-specific population values were obtained from EUROSTAT. The sample weights were trimmed to avoid excessively large sampling variances [46].

Centile curves and reference values were developed using Generalized Additive Models for Location, Scale and Shape (GAMLSS) [47]. Several continuous (Box-Cox Cole and Green (BCCG), Box-Cox power exponential – BCPE, Box-Cox-*t* – BCT, generalized inverse Gaussian) distributions were fitted to the data, optimizing the degrees of freedom (DF) for P-splines fit for all parameters of the respective distributions using Schwarz Bayesian criterion (SBC); appropriate link functions were used for the parameters. BCCG is routinely used in the Lambda Mu Sigma (LMS) method [48]. BCPE and BCT are extensions of LMS adding an extra parameter, *ν*, to allow modelling (positive or negative) kurtosis (with *ν* = 2 BCPE and BCCG (LMS) coincide). In all the models *λ* = 1/3 and *λ* = 1/2 were used for the power transformation of age. Separate analyses were performed for boys and girls. The final model for each test and sex was determined by using SBC. The analysis was performed using R language for statistical computing (R version 3.6.3) [49]; GAMLSS were fitted using R package GAMLSS [50]; post-stratification weights were obtained using R package survey [50]. The best fitting model for each test is presented in **Online Supplementary Table 2**.

## RESULTS

After cleaning and removing outliers, 7,966,693 data points were available, including: 1,026,077 for the 20-m shuttle run; 787,966 for handgrip strength, 1,345,159 for standing long jump, 1,466,821 for body height, 1,466,295 for body mass, 1,464,795 for BMI, and 409,580 for waist circumference. These data came from 106 datasets representing 34 European countries, on children and adolescents aged 6 to 18 years. We originally aimed to collect data as recent as possible to obtain up-to-date reference values, preferably since 2000. Most (69%) datasets (representing 95% of all data points) were collected post-2000, however, pre-2000 data were included when post-2000 were unavailable at the country level. Using these data, we developed CRF and muscular strength reference values (**Tables 1 to 3**) and corresponding percentile curves (**Figure 1**). Reference values for body height, body mass, BMI, and waist circumference are presented in **Online Supplementary Tables 3 to 6**, and **Online Supplementary Figures 1 and 2**. Percentile curves for CRF and muscular strength are higher for boys compared to girls across all ages, with differences increasing with age. The age-related increase in fitness-performance tends to stabilize from age 14 to 15 years onwards. Variation between the fittest (e.g., percentiles 90–99) and least fit (e.g., percentiles 1–10) is larger for boys compared to girls, particularly for the 20-m shuttle run and handgrip strength tests.

**Table 1.**
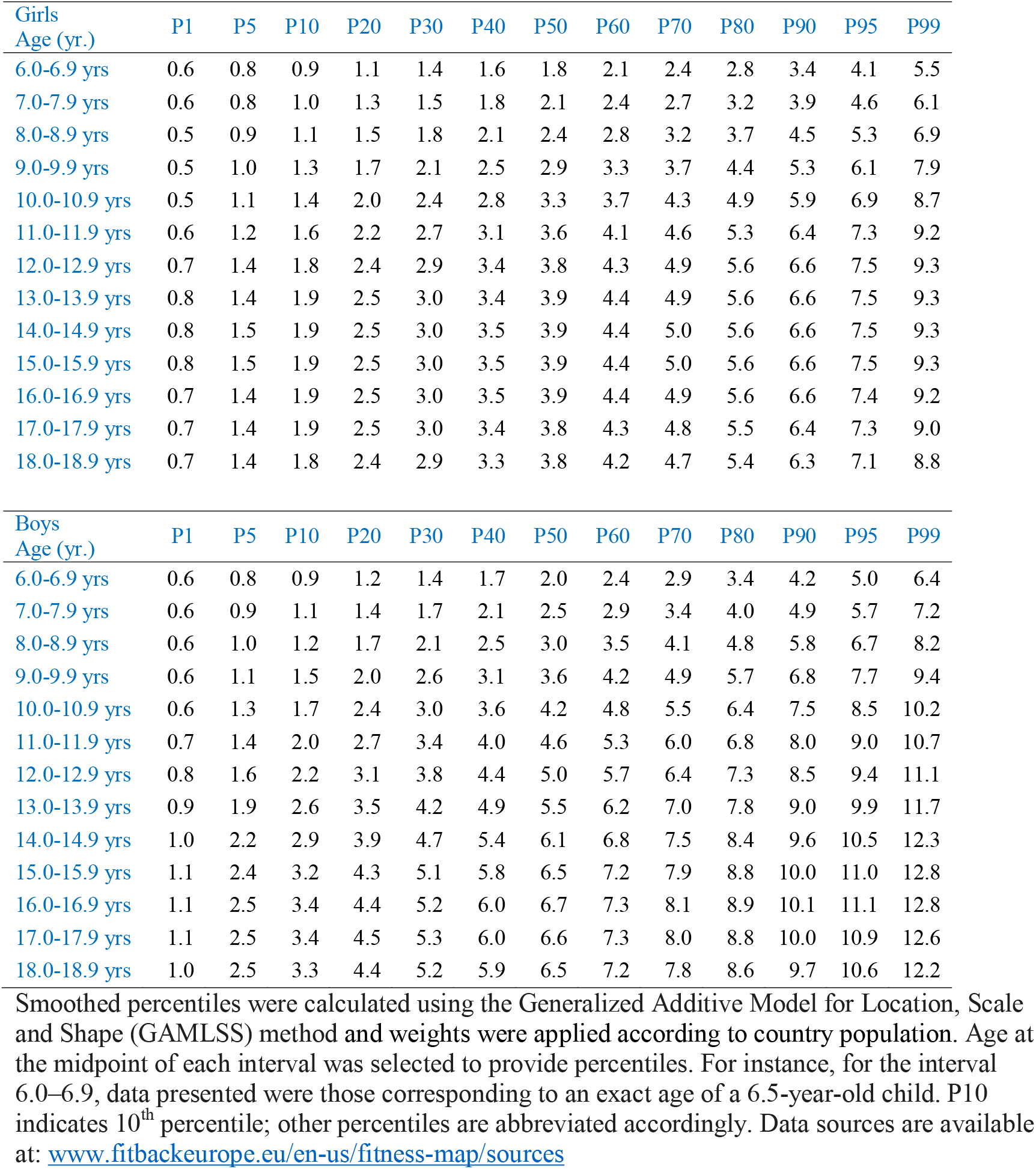
Reference values (centiles) for cardiorespiratory fitness as assessed by the 20-m shuttle run test (expressed in completed stages as a decimal) in European children and adolescents (N=1,063,591)

**Table 2.** Reference values (centiles) for muscular strength as assessed by the handgrip strength test (expressed in kg, average of the maxima for both hands) in European children and adolescents (N=827,585)

| Girls<br>Age (yr.) | P1 | P5 | P10 | P20 | P30 | P40 | P50 | P60 | P70 | P80 | P90 | P95 | P99 |
| --- | --- | --- | --- | --- | --- | --- | --- | --- | --- | --- | --- | --- | --- |
| 6.0-6.9 yrs | 4.2 | 5.6 | 6.3 | 7.2 | 7.8 | 8.3 | 8.8 | 9.3 | 9.9 | 10.7 | 11.8 | 12.9 | 15.5 |
| 7.0-7.9 yrs | 4.7 | 6.4 | 7.3 | 8.4 | 9.1 | 9.7 | 10.4 | 11.0 | 11.7 | 12.6 | 13.9 | 15.3 | 18.4 |
| 8.0-8.9 yrs | 5.2 | 7.4 | 8.5 | 9.8 | 10.7 | 11.4 | 12.2 | 13.0 | 13.8 | 14.9 | 16.6 | 18.1 | 21.9 |
| 9.0-9.9 yrs | 5.8 | 8.4 | 9.7 | 11.3 | 12.3 | 13.3 | 14.1 | 15.1 | 16.1 | 17.3 | 19.3 | 21.1 | 25.6 |
| 10.0-10.9 yrs | 6.7 | 9.7 | 11.3 | 13.0 | 14.3 | 15.3 | 16.3 | 17.4 | 18.5 | 20.0 | 22.2 | 24.3 | 29.3 |
| 11.0-11.9 yrs | 8.0 | 11.6 | 13.3 | 15.3 | 16.8 | 18.0 | 19.1 | 20.3 | 21.6 | 23.2 | 25.7 | 28.1 | 33.7 |
| 12.0-12.9 yrs | 9.5 | 13.6 | 15.5 | 17.8 | 19.3 | 20.7 | 21.9 | 23.2 | 24.6 | 26.4 | 29.1 | 31.6 | 37.8 |
| 13.0-13.9 yrs | 11.1 | 15.5 | 17.7 | 20.0 | 21.7 | 23.1 | 24.5 | 25.8 | 27.3 | 29.2 | 32.0 | 34.7 | 41.1 |
| 14.0-14.9 yrs | 12.4 | 17.1 | 19.3 | 21.8 | 23.5 | 25.0 | 26.4 | 27.8 | 29.3 | 31.2 | 34.1 | 36.9 | 43.4 |
| 15.0-15.9 yrs | 13.1 | 18.0 | 20.2 | 22.8 | 24.6 | 26.0 | 27.4 | 28.8 | 30.4 | 32.3 | 35.2 | 38.0 | 44.6 |
| 16.0-16.9 yrs | 13.4 | 18.4 | 20.8 | 23.4 | 25.1 | 26.6 | 28.0 | 29.4 | 31.0 | 32.9 | 35.8 | 38.6 | 45.2 |
| 17.0-17.9 yrs | 13.7 | 18.9 | 21.3 | 23.9 | 25.7 | 27.2 | 28.6 | 30.0 | 31.5 | 33.4 | 36.3 | 39.1 | 45.7 |
| 18.0-18.9 yrs | 14.3 | 19.6 | 22.0 | 24.6 | 26.4 | 27.9 | 29.2 | 30.6 | 32.2 | 34.1 | 37.0 | 39.7 | 46.3 |

| Boys<br>Age (yr.) | P1 | P5 | P10 | P20 | P30 | P40 | P50 | P60 | P70 | P80 | P90 | P95 | P99 |
| --- | --- | --- | --- | --- | --- | --- | --- | --- | --- | --- | --- | --- | --- |
| 6.0-6.9 yrs | 4.8 | 6.4 | 7.1 | 8.0 | 8.7 | 9.2 | 9.8 | 10.3 | 11.0 | 11.7 | 13.0 | 14.1 | 17.1 |
| 7.0-7.9 yrs | 5.5 | 7.3 | 8.3 | 9.4 | 10.2 | 10.9 | 11.5 | 12.2 | 13.0 | 13.9 | 15.4 | 16.8 | 20.3 |
| 8.0-8.9 yrs | 6.2 | 8.5 | 9.6 | 10.9 | 11.9 | 12.7 | 13.5 | 14.3 | 15.3 | 16.4 | 18.2 | 19.9 | 24.0 |
| 9.0-9.9 yrs | 7.0 | 9.5 | 10.8 | 12.4 | 13.5 | 14.5 | 15.4 | 16.4 | 17.5 | 18.8 | 20.9 | 22.8 | 27.4 |
| 10.0-10.9 yrs | 7.8 | 10.7 | 12.1 | 13.9 | 15.2 | 16.3 | 17.4 | 18.5 | 19.7 | 21.3 | 23.6 | 25.8 | 30.9 |
| 11.0-11.9 yrs | 8.9 | 12.2 | 13.9 | 15.9 | 17.4 | 18.7 | 20.0 | 21.2 | 22.7 | 24.4 | 27.1 | 29.6 | 35.3 |
| 12.0-12.9 yrs | 10.2 | 14.1 | 16.1 | 18.5 | 20.3 | 21.8 | 23.3 | 24.8 | 26.5 | 28.5 | 31.7 | 34.6 | 41.1 |
| 13.0-13.9 yrs | 12.2 | 16.9 | 19.3 | 22.2 | 24.4 | 26.2 | 28.0 | 29.8 | 31.8 | 34.3 | 38.0 | 41.4 | 49.0 |
| 14.0-14.9 yrs | 14.9 | 20.3 | 23.2 | 26.7 | 29.2 | 31.4 | 33.5 | 35.6 | 37.9 | 40.8 | 45.1 | 49.0 | 57.4 |
| 15.0-15.9 yrs | 17.7 | 23.8 | 27.0 | 30.9 | 33.6 | 36.0 | 38.3 | 40.6 | 43.2 | 46.3 | 50.9 | 55.0 | 63.7 |
| 16.0-16.9 yrs | 20.2 | 26.7 | 30.1 | 34.1 | 37.0 | 39.5 | 41.9 | 44.3 | 46.9 | 50.1 | 54.8 | 58.9 | 67.6 |
| 17.0-17.9 yrs | 22.4 | 29.1 | 32.6 | 36.7 | 39.7 | 42.2 | 44.6 | 47.0 | 49.7 | 52.9 | 57.5 | 61.6 | 70.0 |
| 18.0-18.9 yrs | 24.4 | 31.2 | 34.8 | 39.0 | 42.0 | 44.5 | 46.9 | 49.4 | 52.0 | 55.2 | 59.7 | 63.7 | 71.9 |

Smoothed percentiles were calculated using the Generalized Additive Model for Location, Scale and Shape (GAMLSS) method and weights were applied according to country population. Age at the midpoint of each interval was selected to provide percentiles. For instance, for the interval 6.0–6.9, data presented were those corresponding to an exact age of a 6.5-year-old child. P10 indicates 10<sup>th</sup> percentile; other percentiles are abbreviated accordingly. Data sources are available at: www.fitbackeurope.eu/en-us/fitness-map/sources.

**Table 3.**
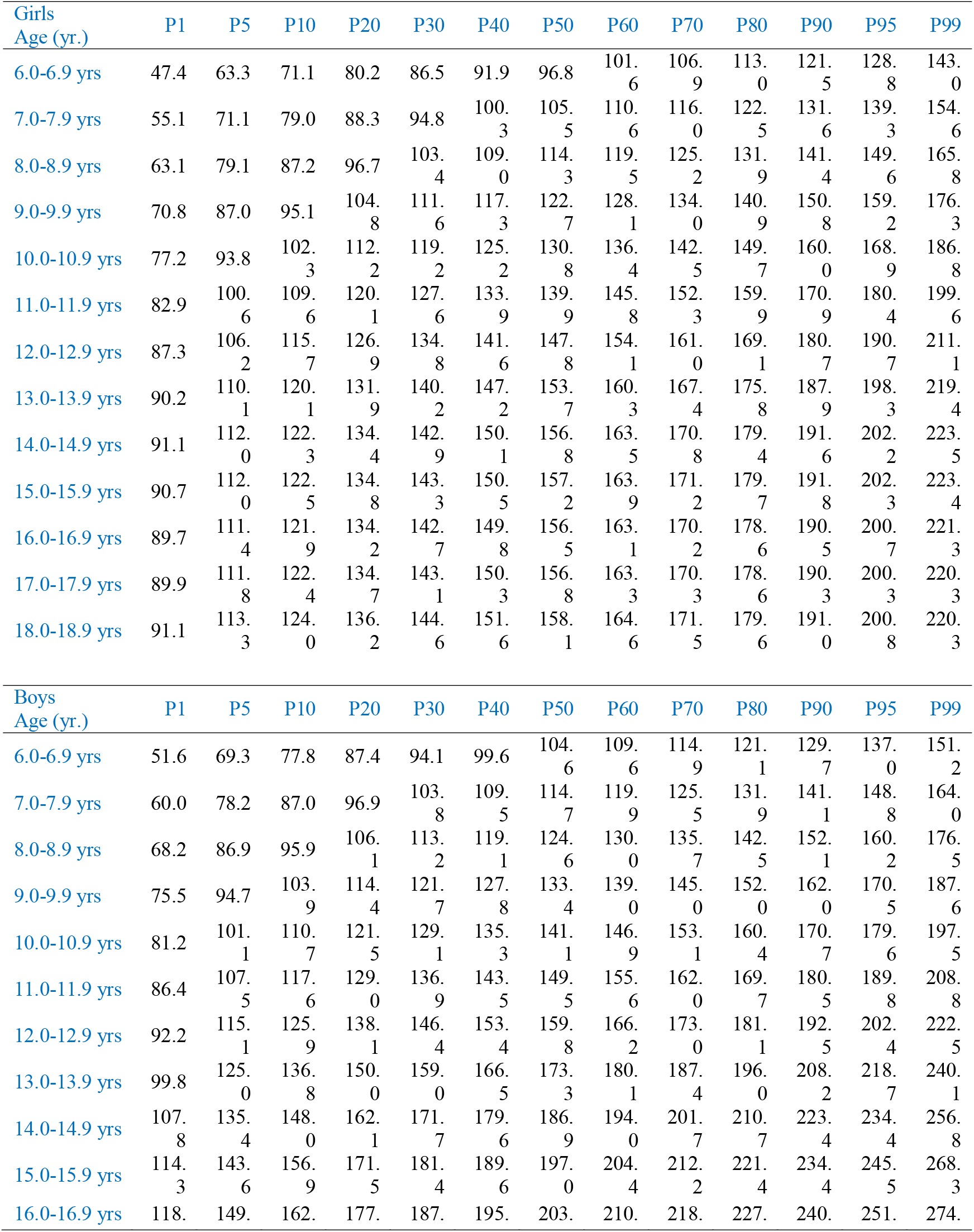
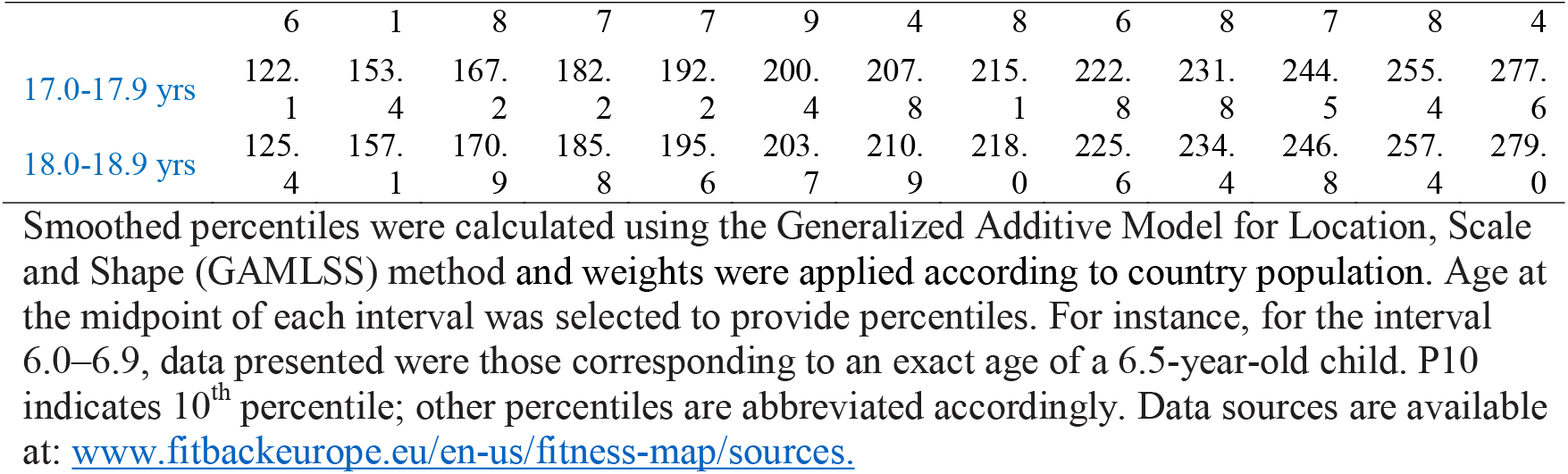
Reference values (centiles) for muscular strength as assessed by the standing long jump test (expressed in cm) in European children and adolescents (N=1,384,856)

**Figure 1.**
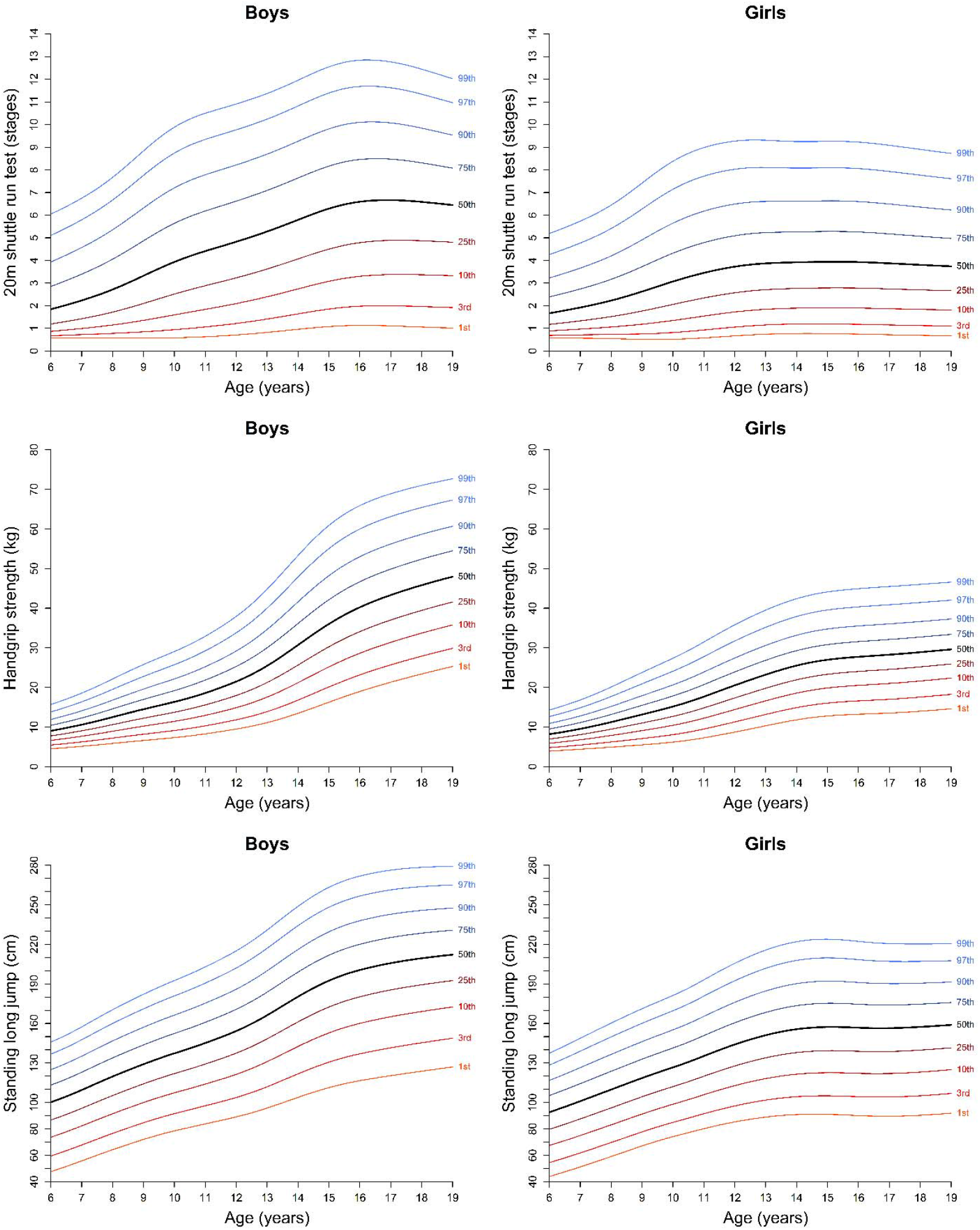
Percentile curves for cardiorespiratory and muscular strength tests among European children and adolescents. Smoothed percentiles were calculated using the Generalized Additive Model for Location, Scale and Shape (GAMLSS) method and weights were applied according to country population. Data sources are available at: https://www.fitbackeurope.eu/en-us/fitness-map/sources.

Mean country-level percentiles and rankings are shown in **Table 4**. Country-level rankings based on mean percentiles are provided for each fitness test, as well as an average estimate for each fitness component (CRF, muscular strength) and the overall European fitness ranking. The top-5 most aerobically fit countries were Iceland, Norway, Slovenia, Denmark, and Finland, and the top-5 physically strong countries were Denmark, Czech Republic, The Netherlands (only one muscular strength test available), Slovenia, and Finland. **Online Supplementary Tables 7 and 8** show the corresponding country-level mean percentile and ranking positions for body height, body mass, BMI, and waist circumference.

**Table 4.**
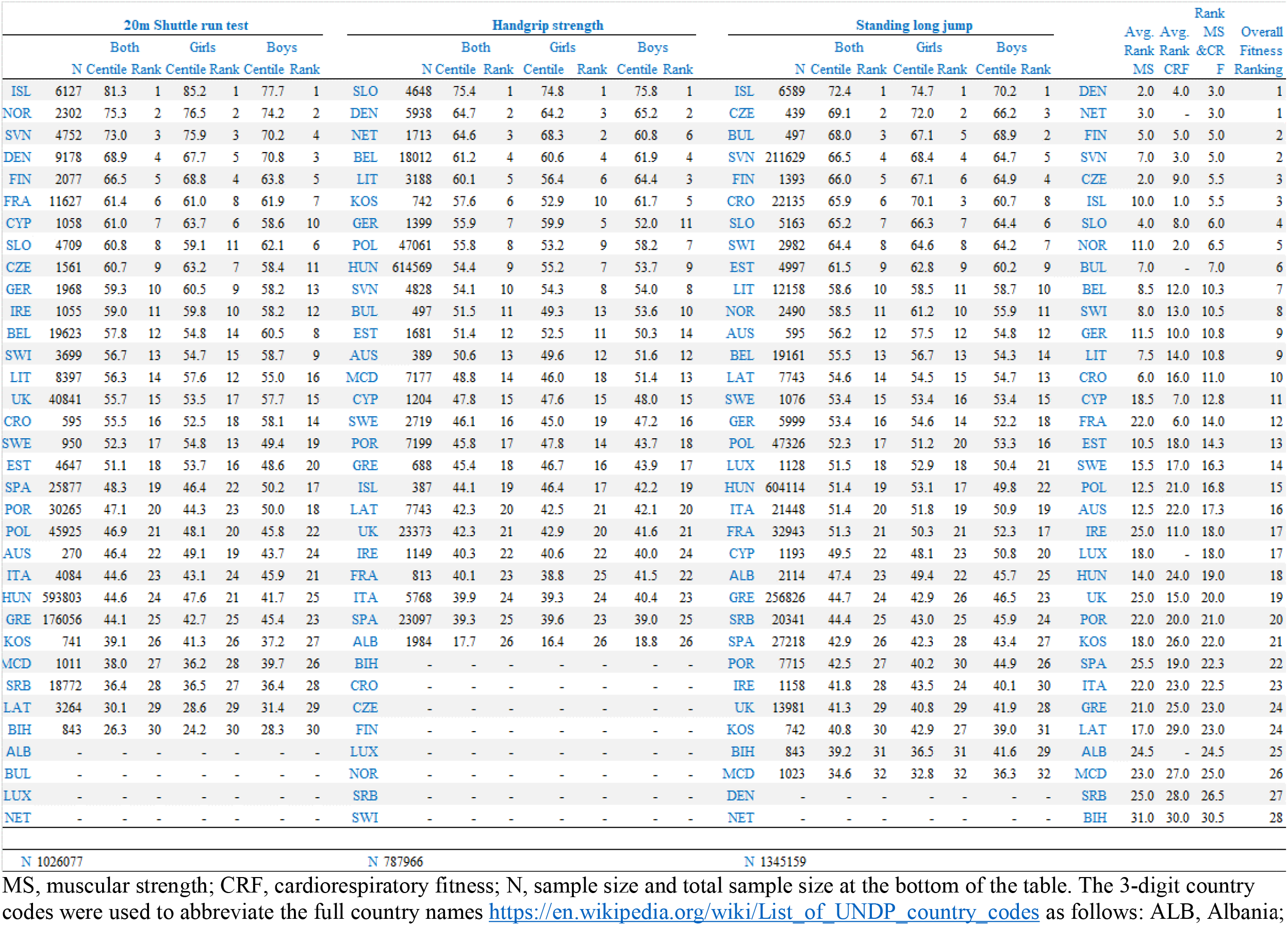
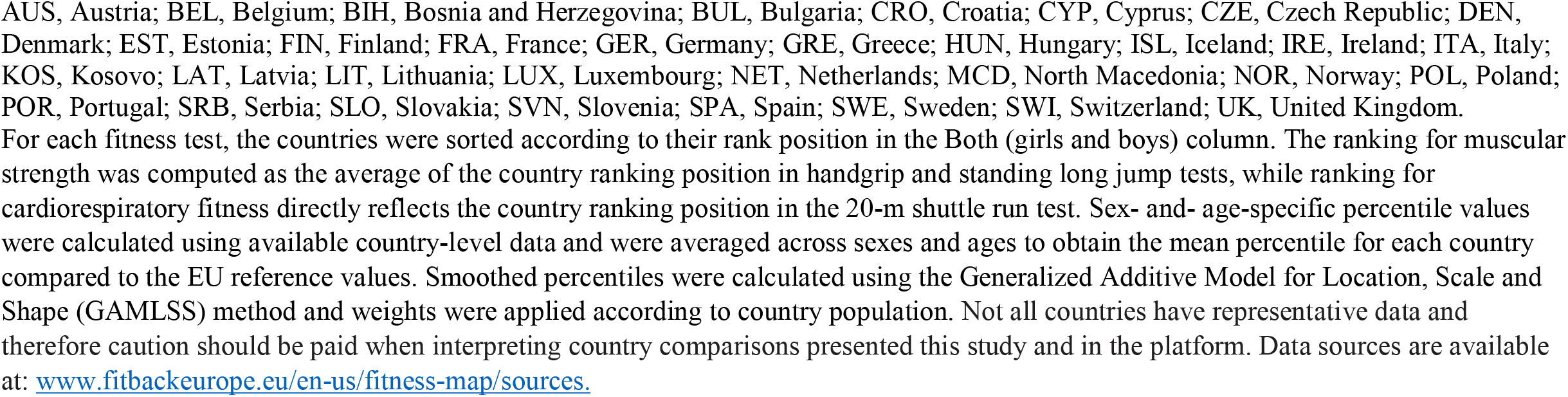
Mean percentile and ranking position of each country according to the pooled EU reference values.

Country comparisons according to mean percentiles are also graphically represented in **Figure 2**, with European fitness maps for each test shown separately. The traffic light color code was used to represent country-specific percentile ranks, with red indicating lower fitness levels, yellow indicating intermediate fitness levels, and green indicating higher fitness levels. The corresponding European maps for BMI and waist circumference are presented as **Online Supplementary Figure 3**. These maps are available in an interactive mode at the FitBack web platform (www.fitbackeurope.eu/en-us/fitness-map) for boys and girls, together and separately. Visual inspection of the fitness maps shows that Southern European countries and the UK generally performed the worst. The correlation between country-level CRF and muscular strength rankings was moderate (r=0.59) and is graphically represented in **Figure 3**. Shaded areas represent those countries ranked in the top-10 for CRF, muscular strength, or both.

**Figure 2.**
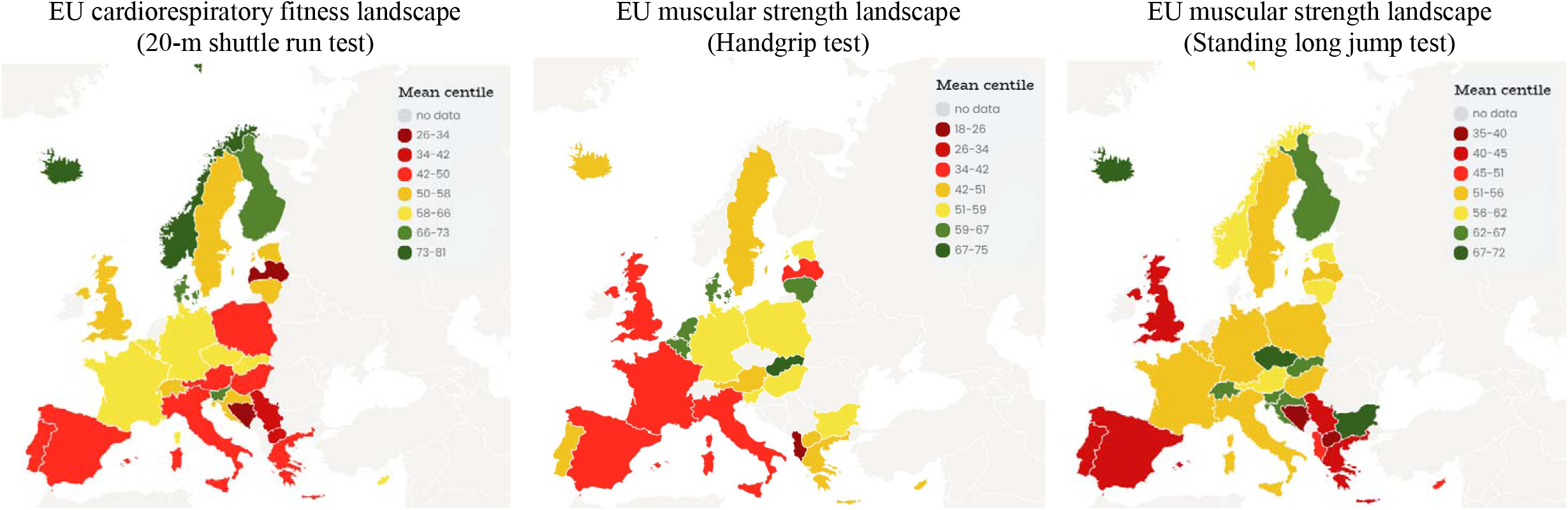
European fitness maps for cardiorespiratory and muscular strength in children and adolescents. Sex- and- age-specific percentile values were calculated using available country-level data and were averaged across sexes and ages to obtain the mean percentile for each country compared to the EU reference values. Smoothed percentiles were calculated using the Generalized Additive Model for Location, Scale and Shape (GAMLSS) method and weights were applied according to country population. Separate European fitness maps for girls and boys for these tests (as well as those for the obesity markers of body mass index and waist circumference) are available at: www.fitbackeurope.eu/en-us/fitness-map. The website map is interactive so that detailed information for each country is shown with the mouseover function. Not all countries have representative data and therefore caution should be paid when interpreting country comparisons presented this study and in the platform. Data sources are available at: www.fitbackeurope.eu/en-us/fitness-map/sources.

**Figure 3.**
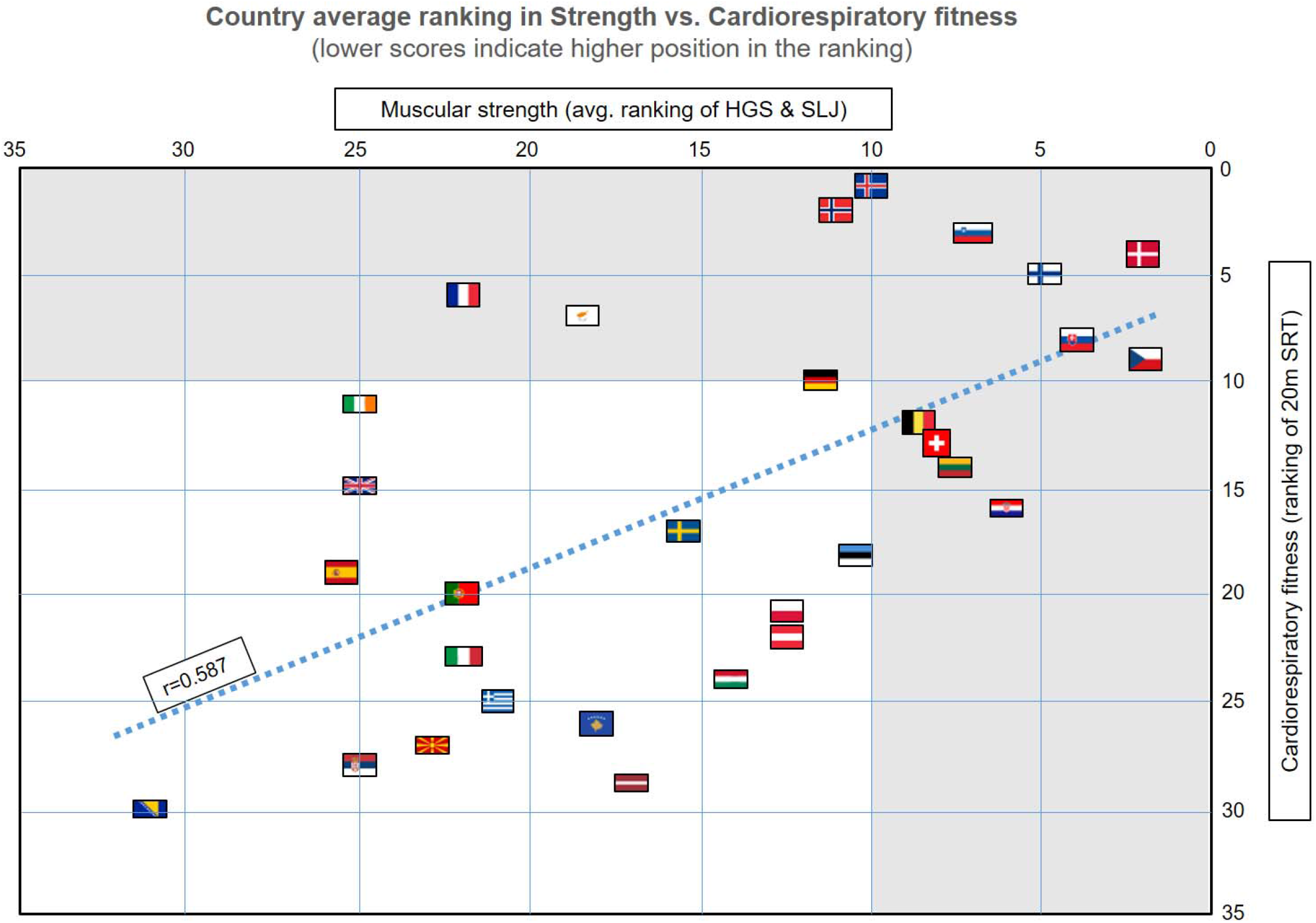
Country average ranking in muscular strength and cardiorespiratory fitness in European children and adolescents. HGS indicates handgrip strength test; SLJ, standing long jump test; 20mSRT, 20-m shuttle run test. The ranking for muscular strength was computed as the average of the country ranking position in handgrip and standing long jump tests, while ranking for cardiorespiratory fitness directly reflects the country ranking position in the 20-m shuttle run test. Gray shaded areas indicate countries ranked in the top-10 for either muscular strength, cardiorespiratory fitness, or both. This figure was created based on the data presented in Table 4. Four countries (Albania, Bulgaria, Luxembourg, and The Netherlands) were not included since they had in either missing muscular strength or cardiorespiratory fitness data. Not all countries have representative data and therefore caution should be paid when interpreting country comparisons presented this study and in the platform. Data sources are available at: www.fitbackeurope.eu/en-us/fitness-map/sources.

## DISCUSSION

### Summary of findings

This article provides the most up-to-date and comprehensive reference values for the health-related fitness of European children and adolescents aged 6–18 years. We also provided country-level mean percentiles for each fitness component. Our overall country-level fitness rankings suggest that Northern (Denmark, Finland, Iceland, and Norway) and Central European countries (Slovenia, Czech Republic, and Slovakia) have the fittest children and adolescents, while Southern European countries (Spain, Italy, and Greece) and the UK are comparatively less fit. Interestingly, we observed a moderate positive correlation between country-level CRF and muscular strength, indicating that despite being different fitness components, children with higher CRF levels generally had higher muscular strength levels. A major contribution of our study is that it comes together with the FitBack web platform (www.fitbackeurope.eu), which is free, multilingual (English, Spanish, French, German, and Italian), and ready to be used by researchers and practitioners in physical education, sport and health, as well as by policy makers across Europe. The FitBack platform provides individual and group-based fitness reports supported by educational materials for implementation of fitness monitoring to support fitness education (i.e., to help understand why fitness and fitness testing are important, how to interpret fitness test results, how to set exercise goals, how to improve fitness levels, etc.) and improve physical literacy, as well as interactive European fitness maps based on our reference values. To date, the best available fitness reference values for a large sample of European children and adolescents were those published by Tomkinson et al. in 2018 [39]. Our study updates such work, by expanding the Tomkinson et al. data set [39] and updating the CRF and muscular strength reference values with more recent and representative data for each country.

### Usefulness and practical implications of fitness testing and monitoring

Our reference values, when integrated into the interactive FitBack web platform, have practical utility and implications. First, fitness testing and monitoring is extremely important from a public health and clinical point of view, as recently acknowledged by the American Heart Association [13], and others [51]. Measuring cardiometabolic risk factors from blood samples is invasive and ethically questionable for youth at the population level. Likewise, mental and cognitive health assessments are often complex, sensitive and time consuming. Since physical fitness has repeatedly and consistently been shown to be a powerful marker of physical, mental, and cognitive health in youth, fitness testing and monitoring will provide valuable insight into the health status of youth at individual and group levels. However, clinicians may not have the time, resources, facilities, or expertise to conduct fitness testing (e.g. the 20m shuttle run test) in clinical settings. Therefore, we believe that the most feasible alternative and future goal is that population-level fitness testing be conducted in schools, with test results and interpretation incorporated into the healthcare system databases and forming part of an individual’s medical records that can be viewed by pediatricians and school doctors/nurses. Such practice has been implemented at the regional level in Galicia, Spain [52], and at the national level in Slovenia [40] and Finland [53]. In addition, our article and the interactive FitBack website provide a valuable and cost-effective solution for establishing fitness monitoring at the school, community, regional and national level. For instance, policy makers at education, sport, and health institutions can obtain valuable information about regional differences or temporal trends by monitoring fitness levels over time and use these reference values and the FitBack tool for proper sex- and age-specific interpretation.

In fact, fitness monitoring could flag a sudden decline in fitness, and therefore health, due to unique/unexpected situations, such as COVID-19 pandemic-related lockdowns and the substantial, rapid declines in youth fitness levels reported in countries with fitness surveillance systems [54,55]. Thus, timely interventions for specific target groups can be implemented.

Second, fitness monitoring is part of physical education curricula in many European countries, but most European physical education teachers do not currently have access to an easy-to-use and automatic tool for interpreting sex- and age-specific fitness test results. With our article and the FitBack platform, we aimed to contribute to an extensive implementation of fitness monitoring across European schools. In this context, the FitBack platform also provides information to avoid undesirable practices, such as grading students based on their fitness levels and fitness competitions among students, by using fitness testing as an educational tool to facilitate learning and understanding about fitness and its importance to health and sport, and setting individual goals for improvement. Such an approach to fitness testing should help improve physical literacy among European youth. Enhancing physical fitness through goal setting and an appropriate physical activity program, and tracking changes through fitness monitoring, may improve students’ physical literacy journey. Those with better fitness education may be more attuned with their body and what is required to function well, and may be able to foster lifelong physical activity habits.

Third, our reference values can be used for sport/athletic profiling and monitoring, as well as talent identification and development [42,56]. Youth who have fitness levels above the 90^th^percentiles may be considered talented in certain fitness components and sports participation could be promoted to them and their family. Likewise, changes in fitness levels in response to a lifestyle intervention could be tracked against our sex- and age-specific percentile bands to identify expected, better than expected, or worse than expected developmental changes.

### Limitation and strengths

While the FitBack network gathered 8 million data points for the development of new health-related reference values, the included data are not representative of all European youth. Some countries such as Slovenia, Hungary, and Portugal (www.fitbackeurope.eu/en-us/monitoring-fitness/best-practice) have established fitness monitoring systems that cover all school-age youth. Other countries such as Greece [57] and Poland [58] have conducted nationally-representative fitness testing at particular points in time, while most European countries do not have nationally-representative fitness data available. This implies that our country-level comparisons should be taken cautiously given that not all data are representative of their source populations. Our ambition was to identify the best available and most recent data (using the ALPHA fitness tests) for each country to update existing CRF and muscular strength reference values, and to strengthen the evidence supporting the FitBack platform. Important contributions from our study and the FitBack network include: (1) increased awareness around the importance of fitness surveillance and monitoring, (2) the identification of countries that have access to large fitness databases, and (3) to facilitate fitness testing and interpretation through the FitBack platform, which we hope will improve the amount, quality, and availability of future fitness data. Unfortunately, included fitness data were collected at different times and temporal trends in fitness may have biased our results. To minimize the potential for bias, old data collected in 1980s were excluded from our analyses, with 95% of our data points collected since 2000 (see Online Supplementary Table 1). Only harmonized cross-country testing at the same time will provide the most accurate comparisons. While not nationally representative, the HELENA study collected harmonized fitness data in 2005–08 across 10 European cities, and the results suggested that adolescents living in Southern Europe (Spain, Italy, Greece) had lower levels of CRF and muscular strength, as well as more total and central adiposity, than their peers living in Central-Northern Europe [59]. These findings are consistent with the FitBack results hereby presented, and are in line with previous reports[60,61]. Another limitation of our study is the protocol variation across studies. In order to improve this moving forward, we recommend researchers use the ALPHA fitness test battery manuals of operations and explanatory videos that are freely available (http://profith.ugr.es/alpha-children available in English and Spanish), and which have been incorporated into the FitBack platform (www.fitbackeurope.eu/en-us/make-report/about-testing). Finally, while we obtained data from 77% (34/44) of European countries (https://www.schengenvisainfo.com/countries-in-europe/), additional data are required from the remaining countries to paint a complete European fitness picture.

## Conclusion

There is overwhelming evidence supporting the importance of fitness testing from a health, educational, and sport point of view. Further, the EU-funded ALPHA project identified the most reliable and valid fitness tests, providing the methods (manuals of operations, videos) needed to evaluate youth health-related fitness levels in a standardized manner across Europe. Now, the FitBack project provides the scientific and practitioner communities with the steps needed for the implementation of youth-based fitness assessment and interpretation in school or sporting settings across Europe. Our sex- and age-specific reference values have practical implications and are the foundation of the FitBack platform for interactive individual and group-based interpretation of fitness levels. These reference values should be revisited in the future as more countries introduce national surveillance systems to reflect the updated fitness levels of European youth. The FitBack network, therefore, welcomes new members and is searching for missing and new fitness data.

## Data Availability

The availability of each of the datasets used in this study needs to be requested separately to the authors owning their data, which can be found in the references cited in the Online Supplementary Table 1.

## Acknowledgements

The authors acknowledge the support of all FitBack network members who provided data on physical fitness of children and adolescents (see: www.fitbackeurope.eu/en-us/fitness-map/sources) including large EU-funded consortium projects such as HELENA and IDEFICS. We thank D. Mayorga-Vega for his assistance on obtaining the fitness data from Poland.

## ONLINE SUPPLEMENTARY MATERIAL

**Online Supplementary Table 1.**
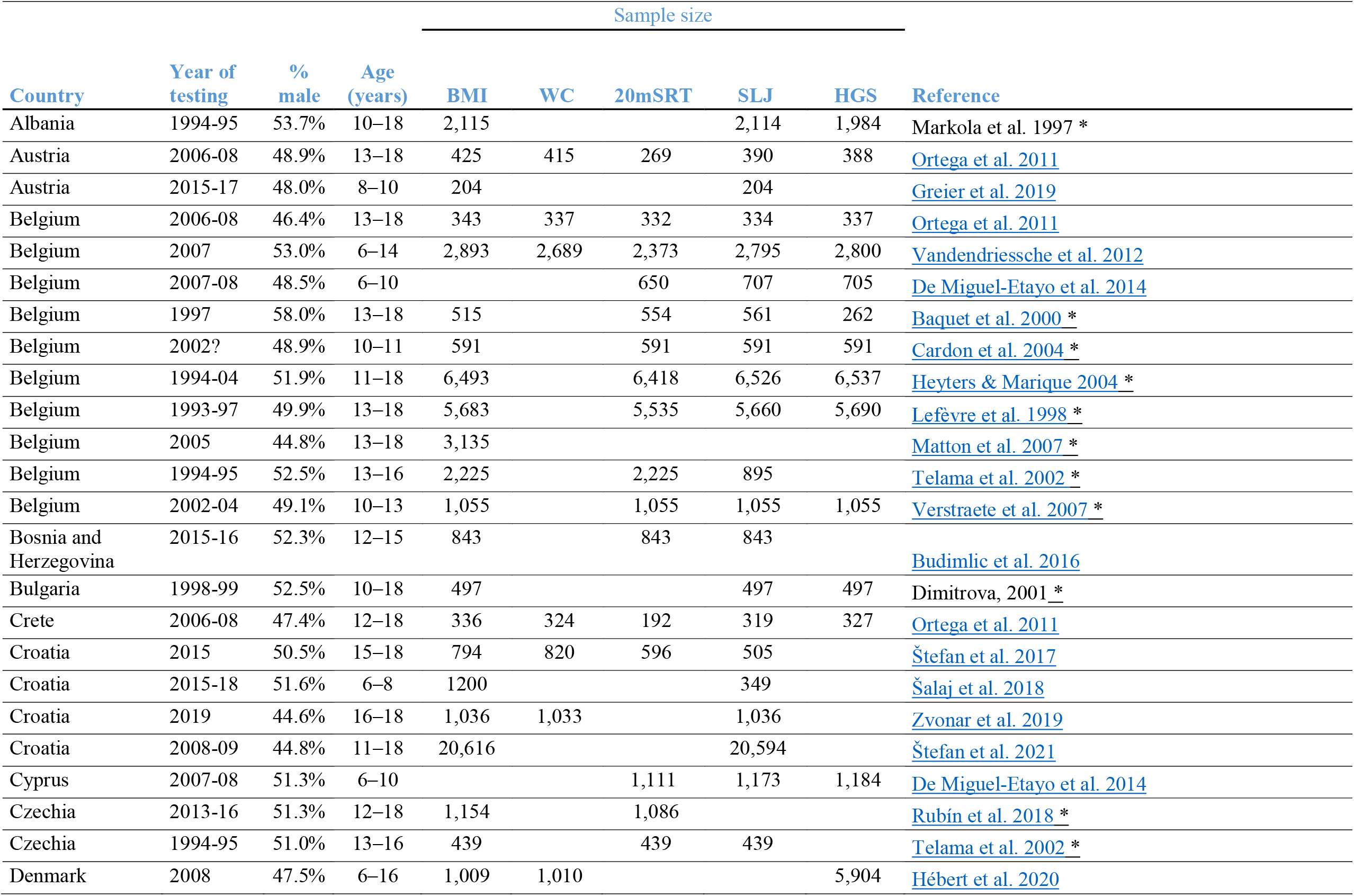
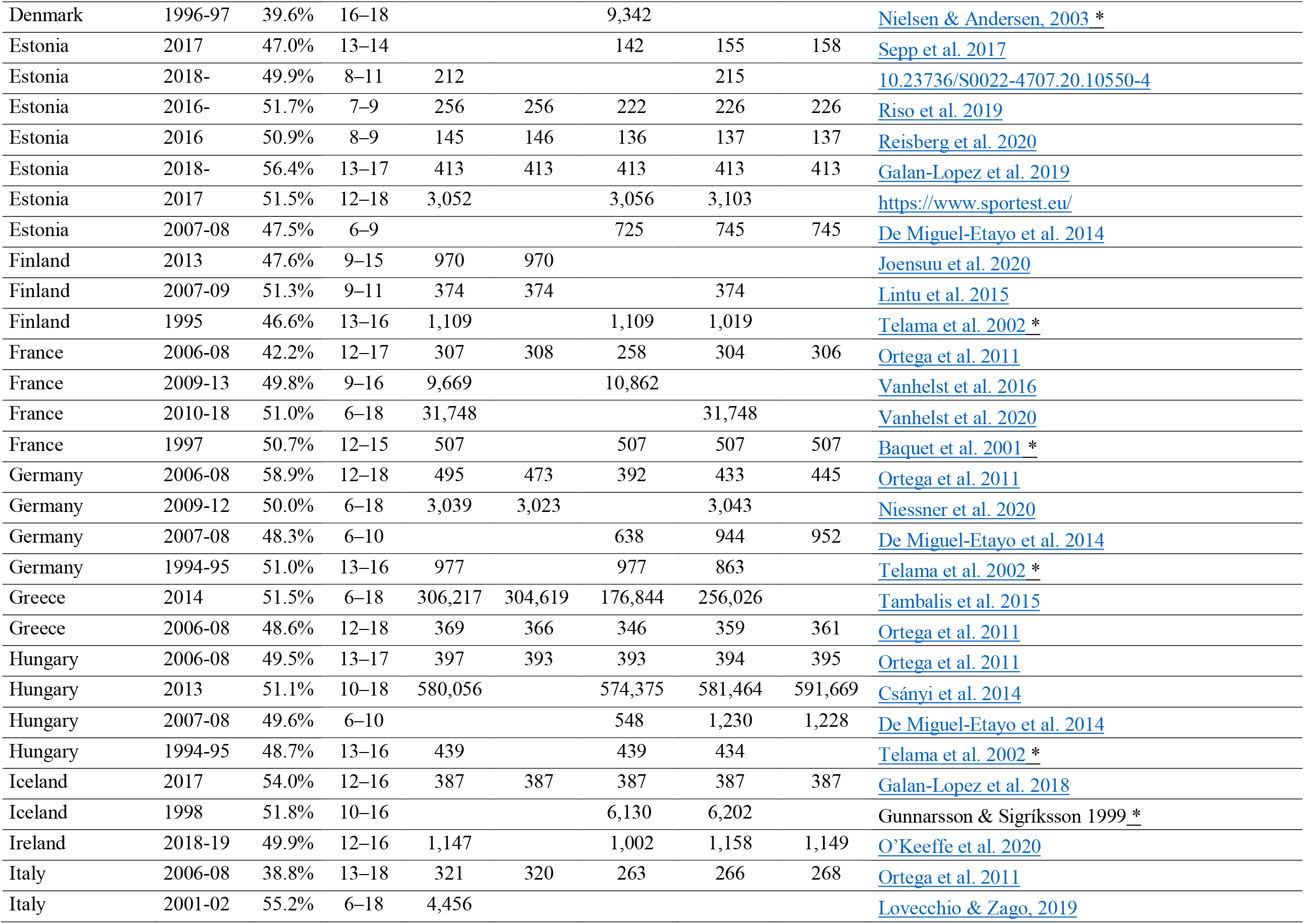
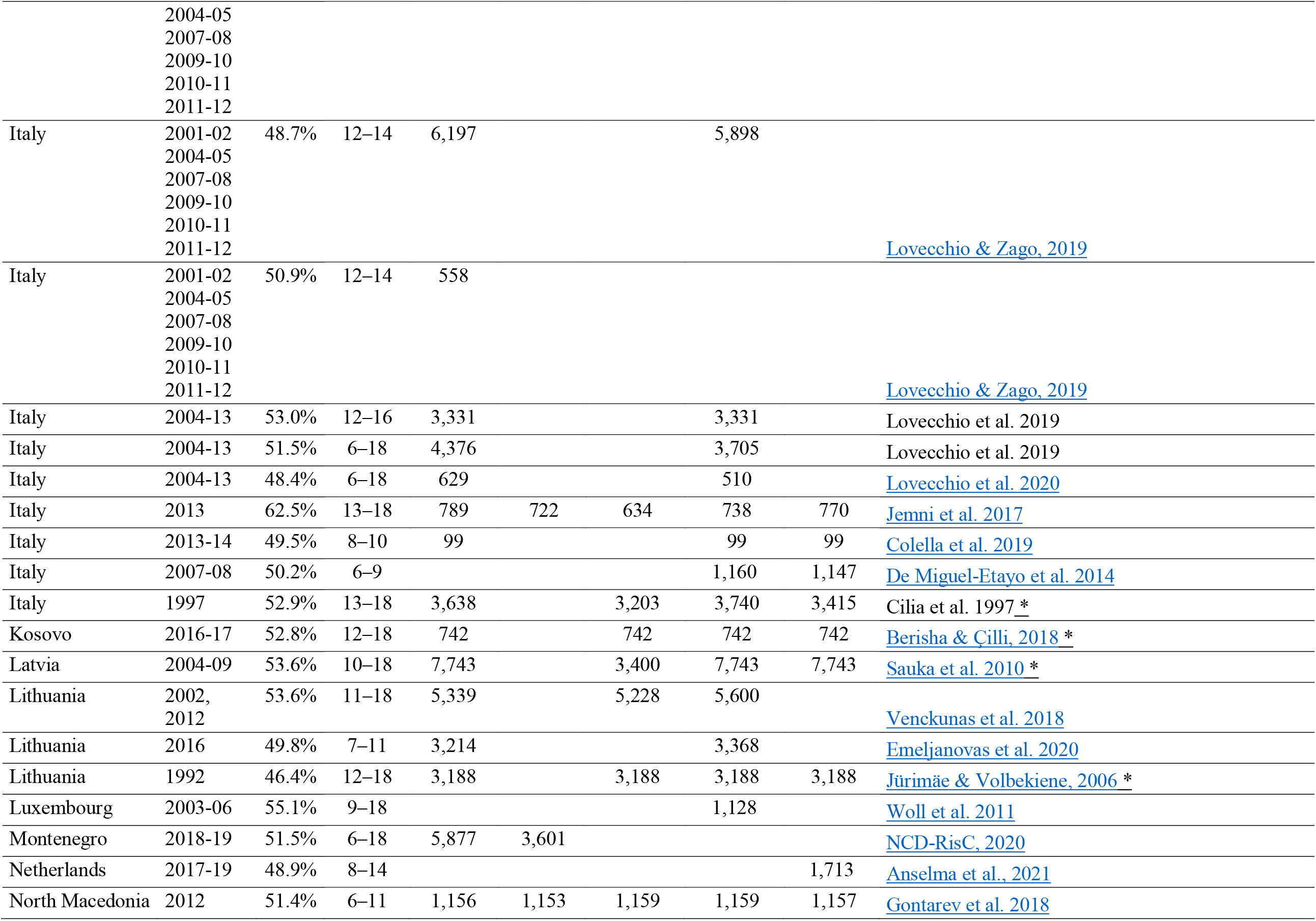
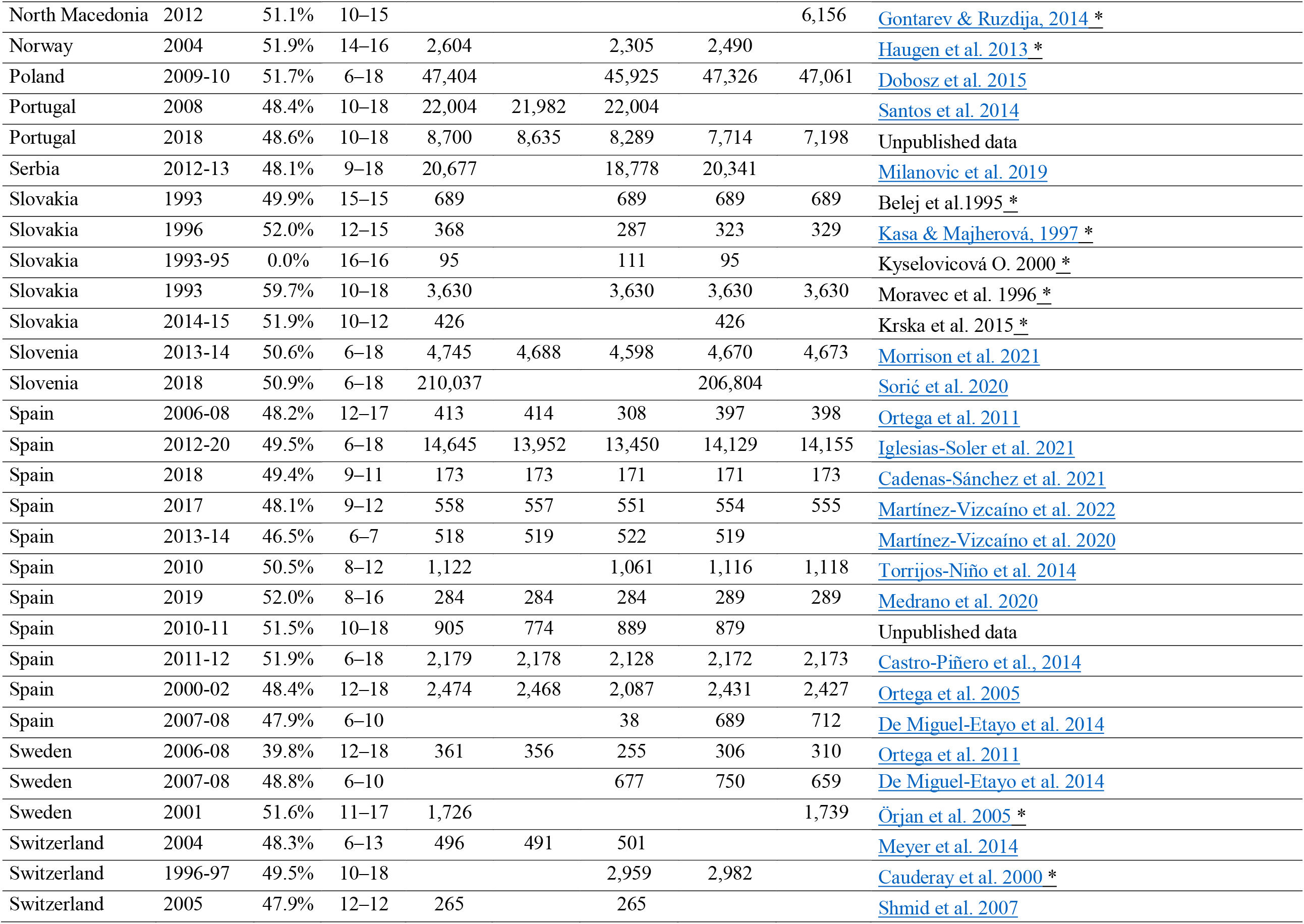
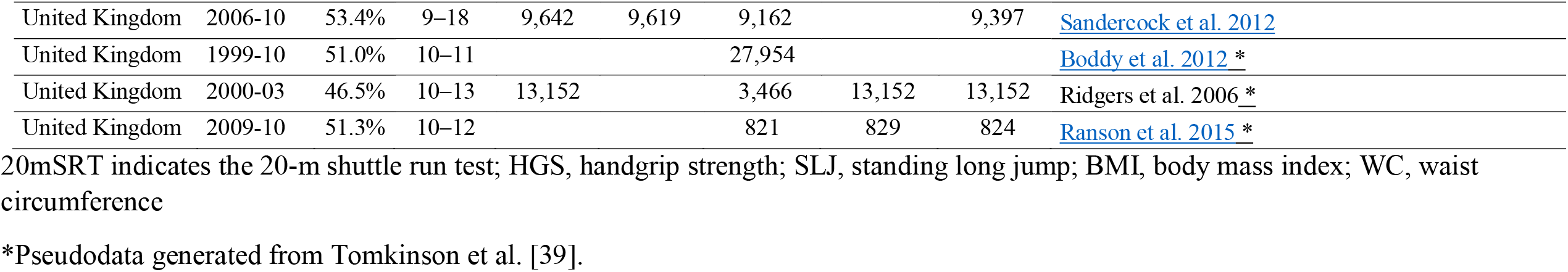
Summary of the sources used for generating the FitBack reference values and centiles.

**Online Supplementary Table 2.**
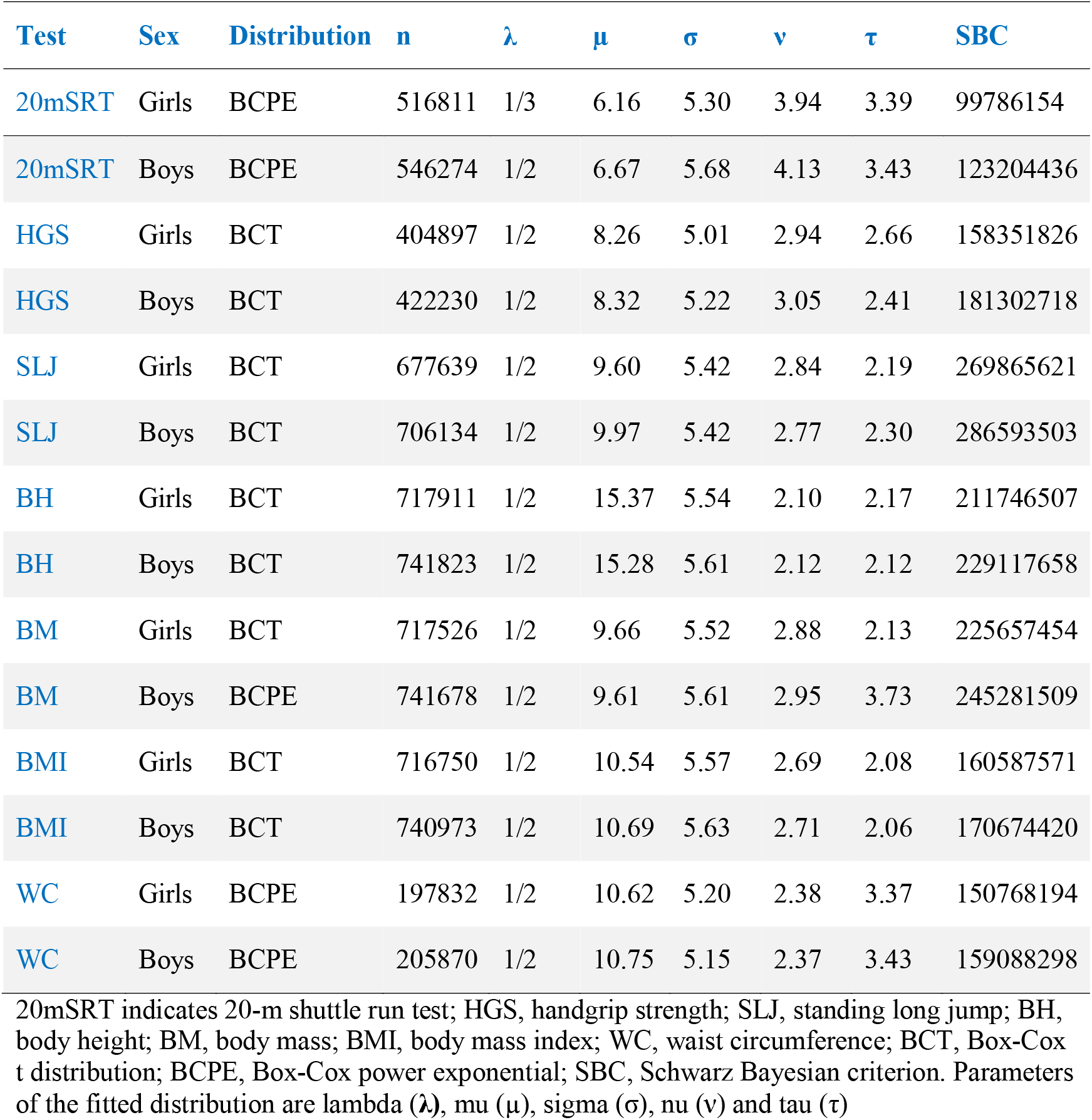
Generalized Additive Model for Location, Scale and Shape (GAMLSS) models used to calculate the physical fitness smoothed percentiles.

**Online Supplementary Table 3.**
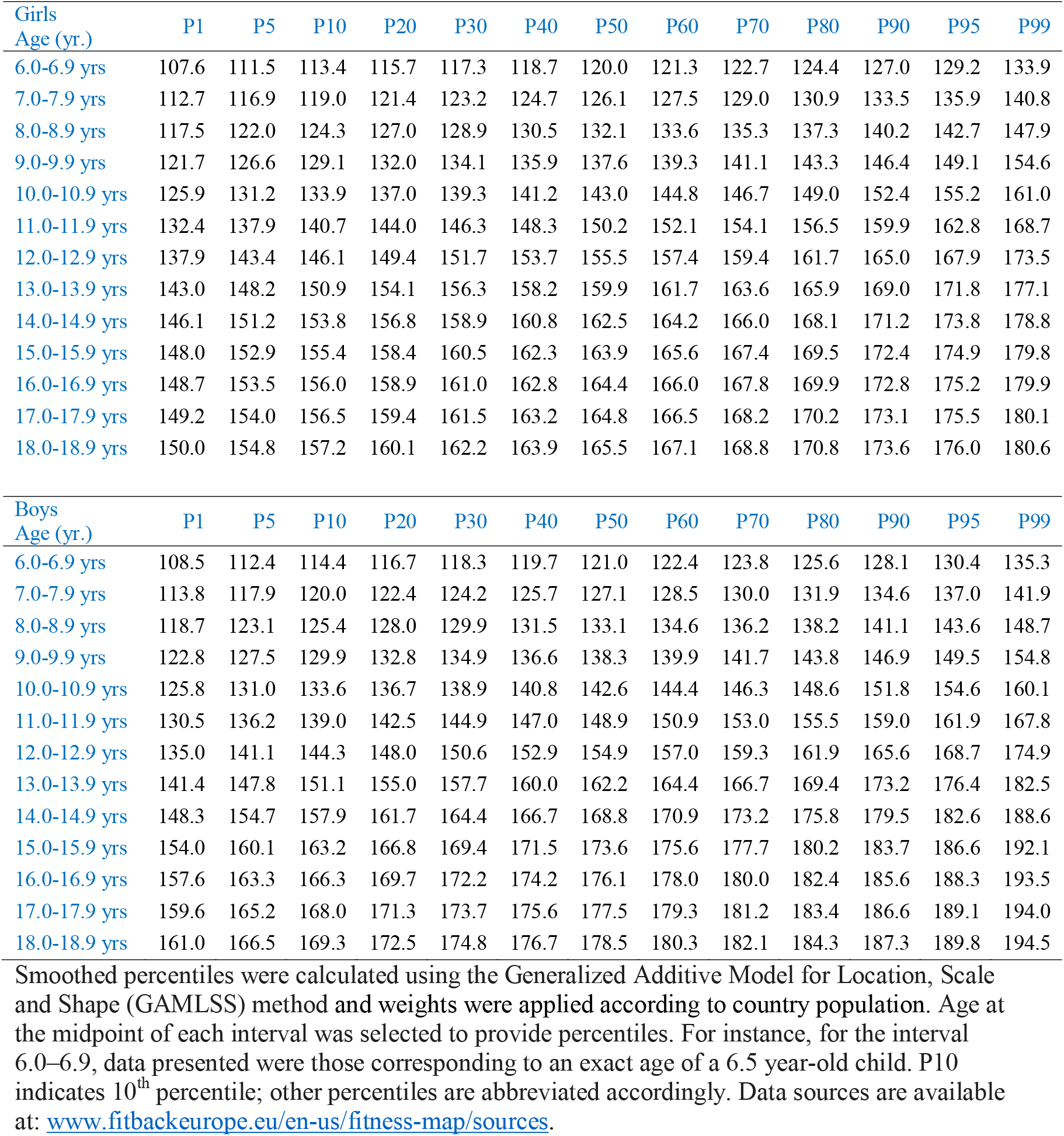
Reference values (centiles) for body height (cm) in European children and adolescents (N=1,466,821)

**Online Supplementary Table 4.**
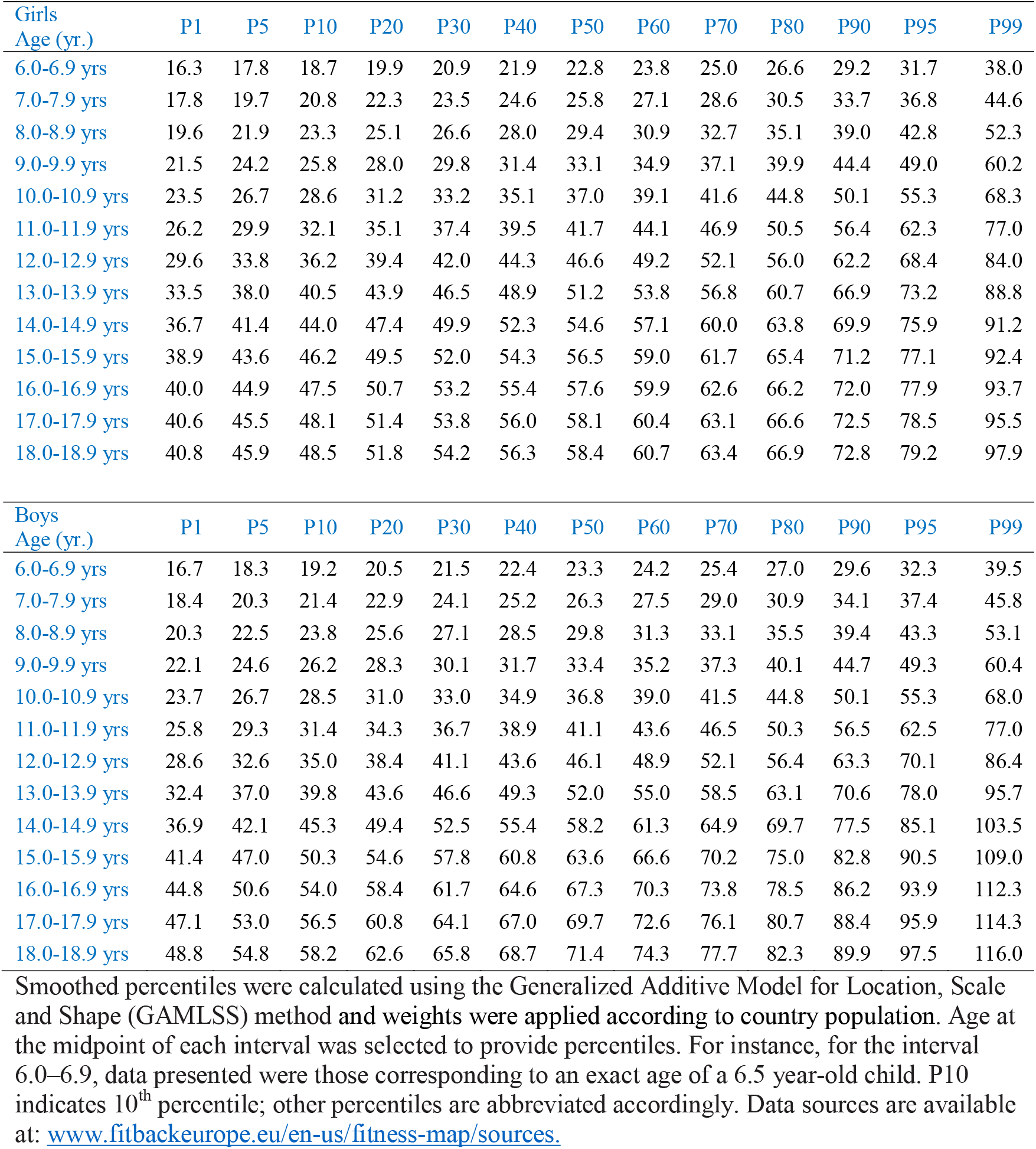
Reference values (centiles) for body mass (kg) in European children and adolescents (N=1,466,295)

**Online Supplementary Table 5.**
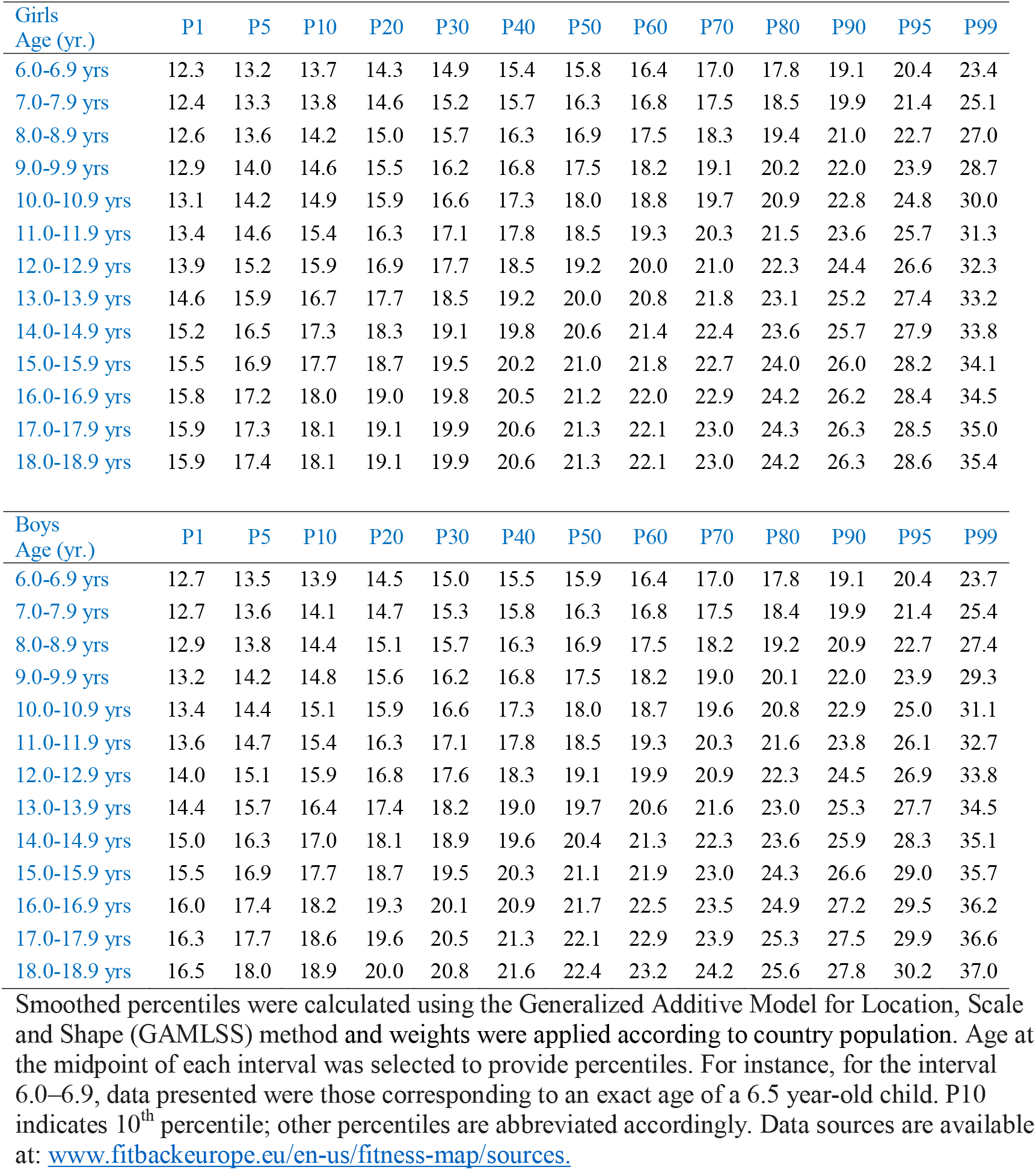
Reference values (centiles) for body mass index (kg/m^2^) in European children and adolescents (N=1,464,795)

**Online Supplementary Table 6.**
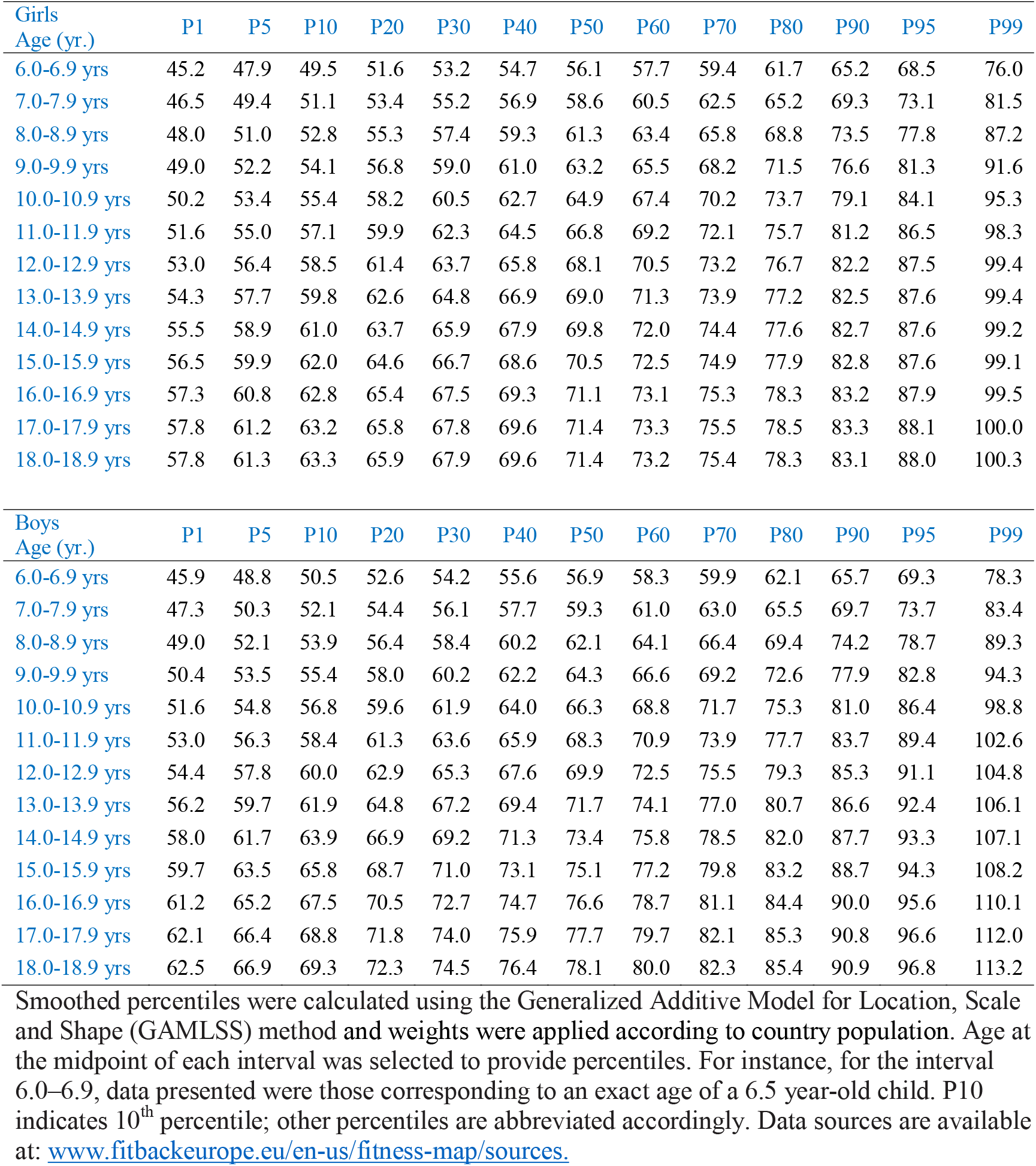
Reference values (centiles) for waist circumference (cm) in European children and adolescents (N=409,580)

**Online Supplementary Table 7.**
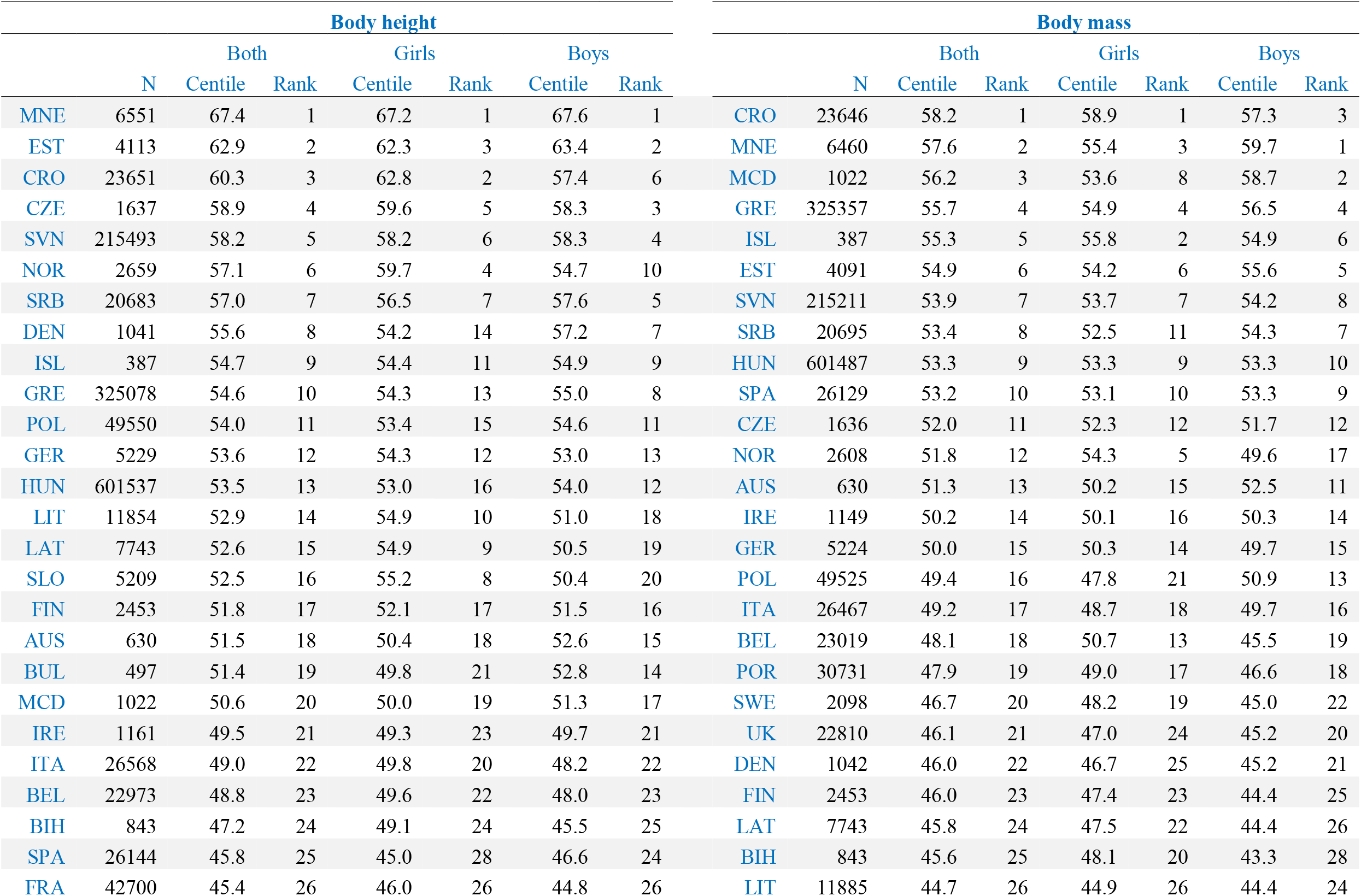
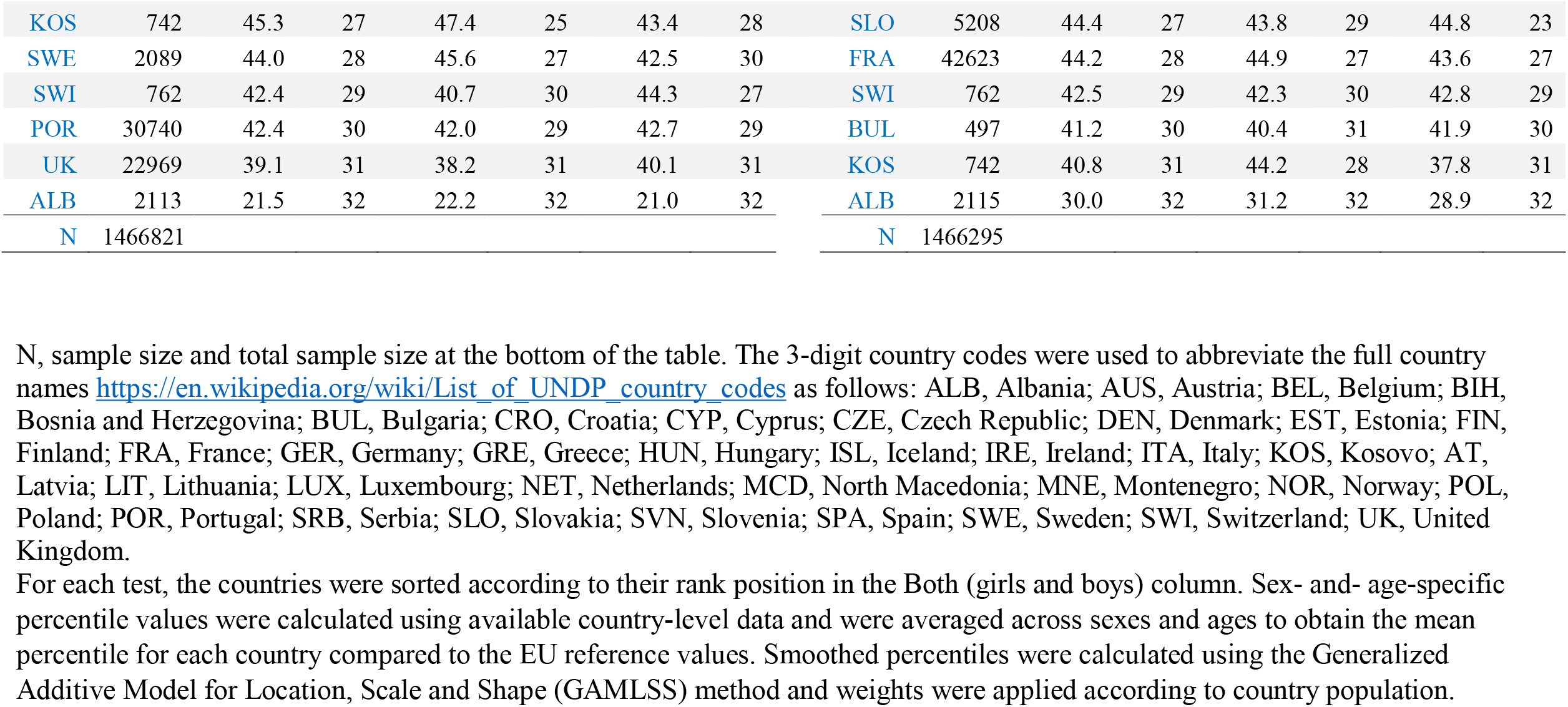
Mean percentile and ranking position of each country according to the pooled EU reference values for body height and weight.

**Online Supplementary Table 8.**
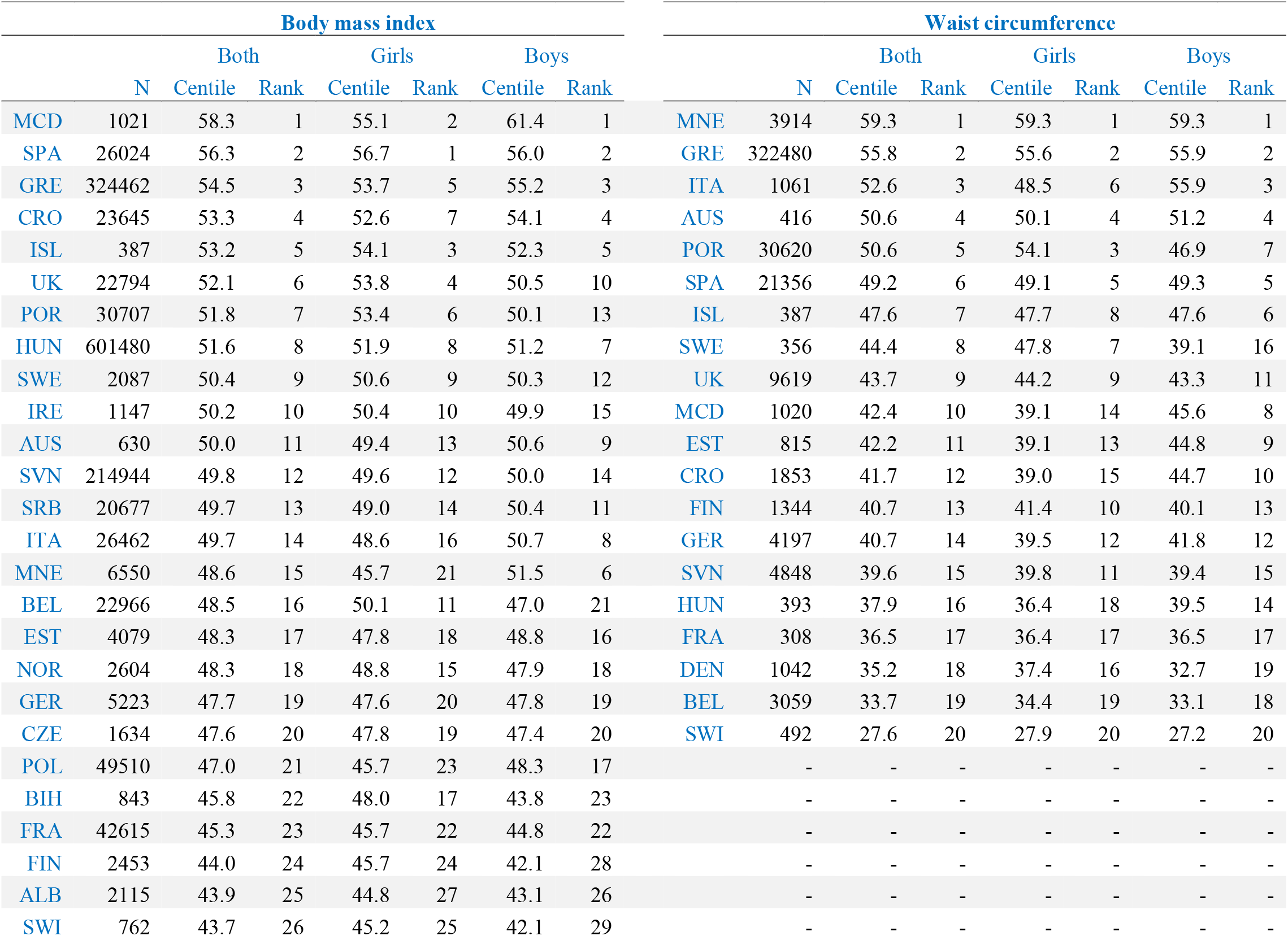
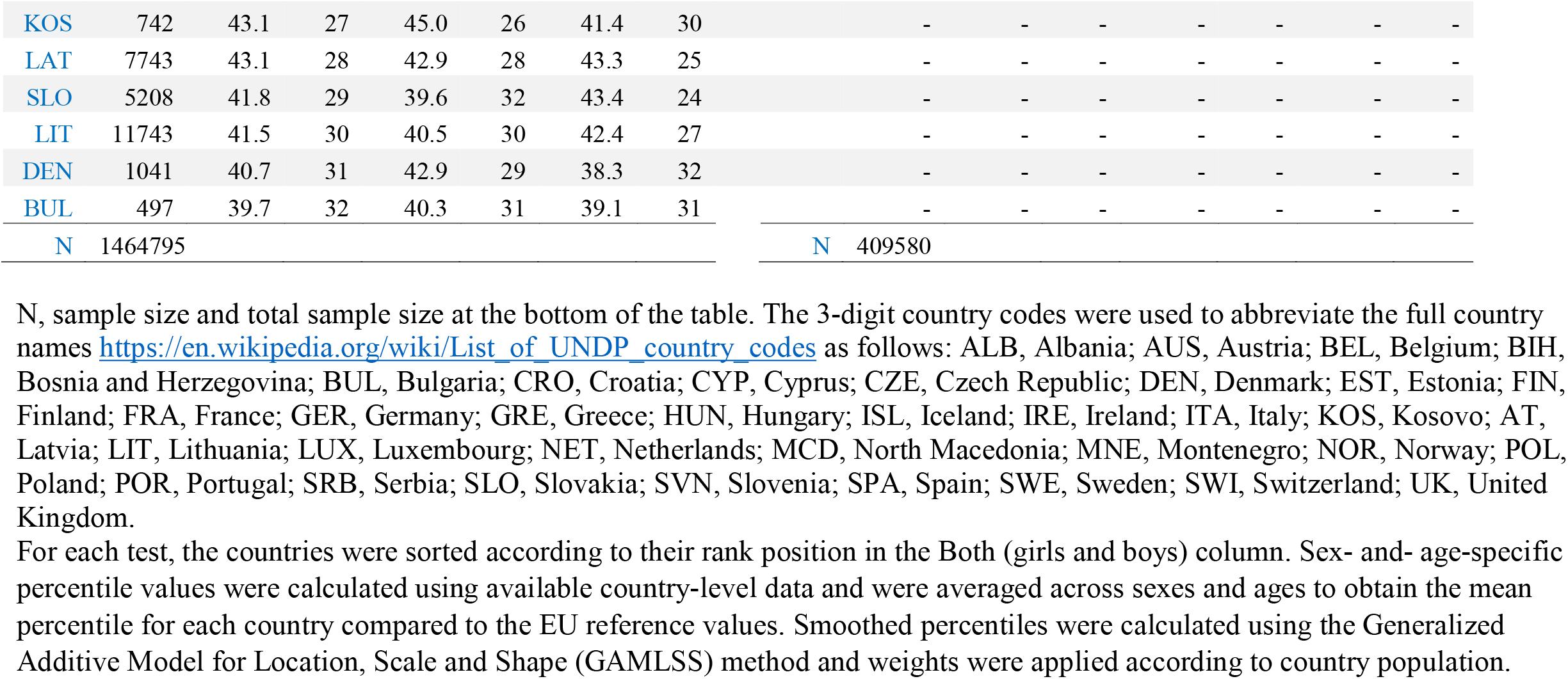
Mean percentile and ranking position of each country according to the pooled EU reference values for body mass index and waist circumference.

**Online Supplementary Figure 1.**
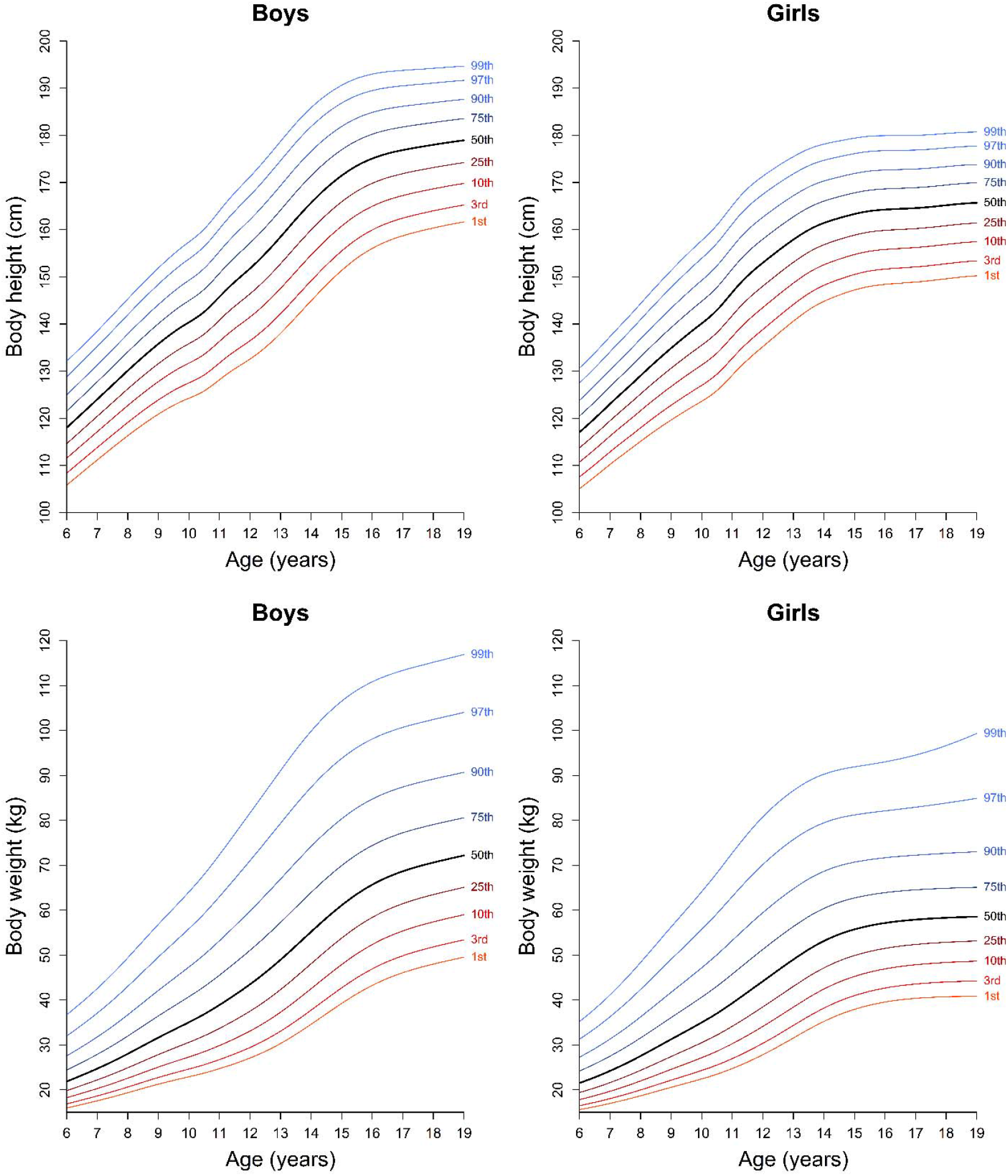
Percentile curves for body height and body mass in European children and adolescents. Smoothed percentiles were calculated using the Generalized Additive Model for Location, Scale and Shape (GAMLSS) method and weights were applied according to country population. Data sources are available at: https://www.fitbackeurope.eu/en-us/fitness-map/sources.

**Online Supplementary Figure 2.**
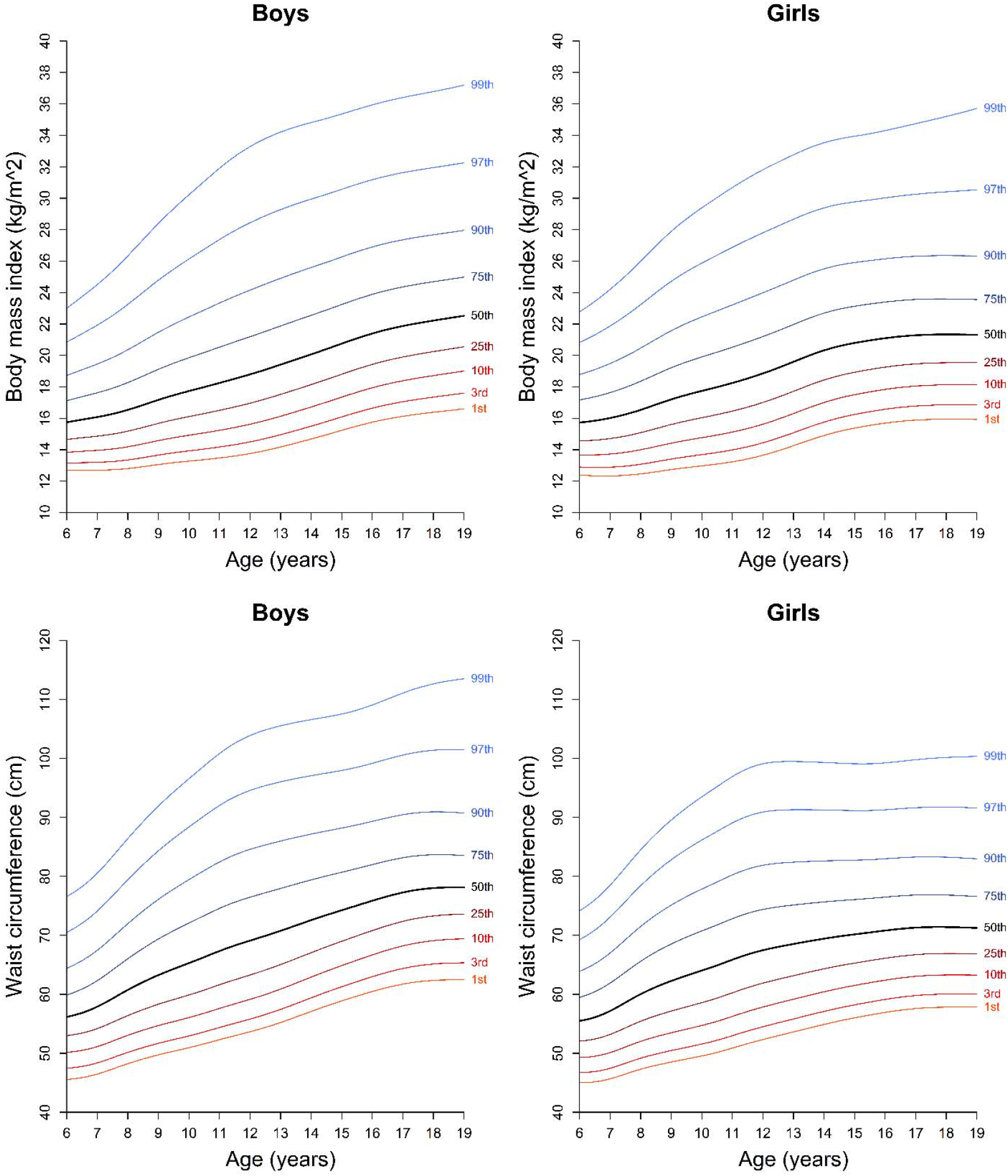
Percentile curves for body mass index and waist circumference in European children and adolescents. Smoothed percentiles were calculated using the Generalized Additive Model for Location, Scale and Shape (GAMLSS) method and weights were applied according to country population. Data sources are available at: https://www.fitbackeurope.eu/en-us/fitness-map/sources. EU body mass index landscape EU waist circumference landscape

**Online Supplementary Figure 3.**
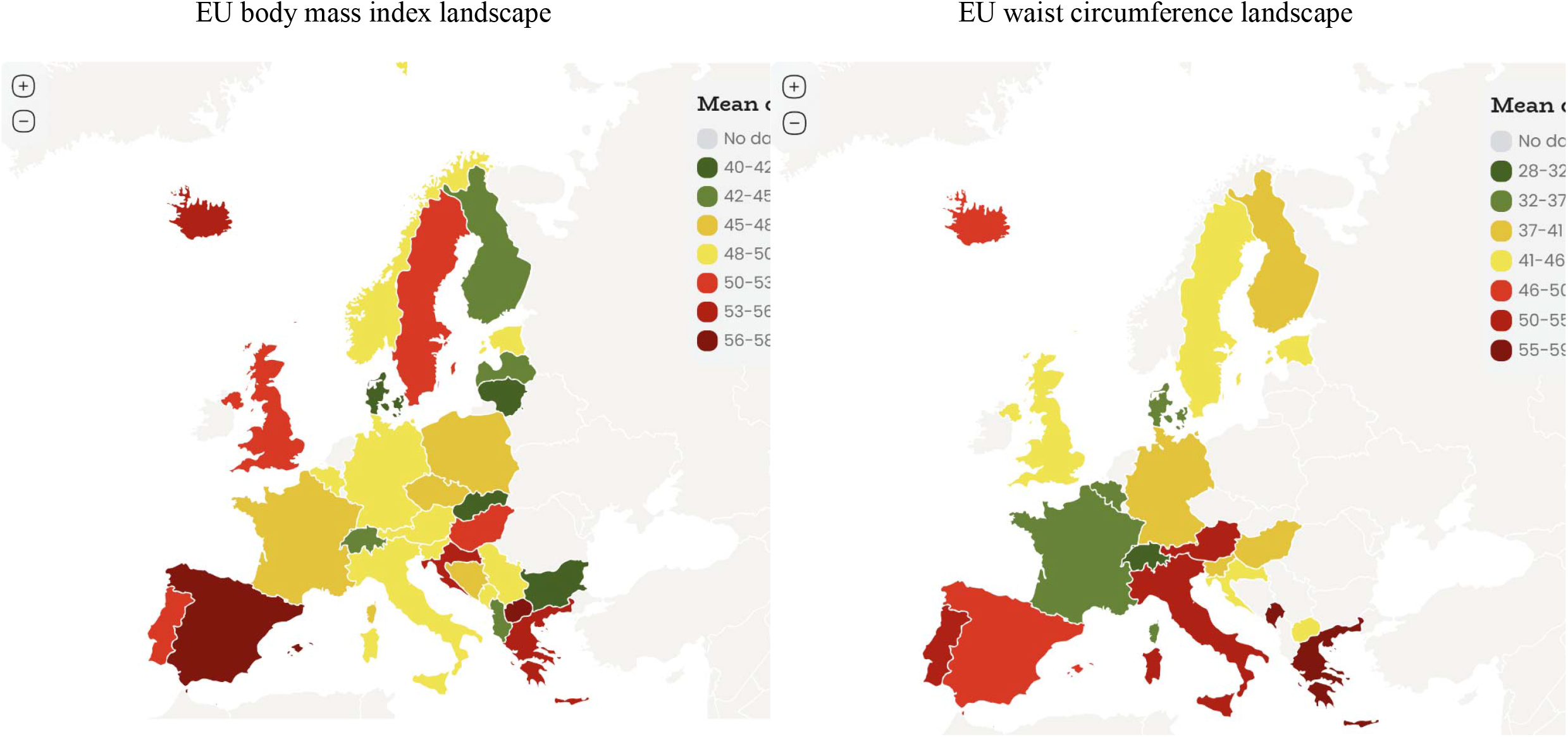
European maps for body mass index and waist circumference in children and adolescents. Sex- and- age-specific percentile values were calculated using available country-level data and were averaged across sexes and ages to obtain the mean percentile for each country compared to the EU reference values. Smoothed percentiles were calculated using the Generalized Additive Model for Location, Scale and Shape (GAMLSS) method and weights were applied according to country population. Separate European fitness maps for girls and boys are available at: www.fitbackeurope.eu/en-us/fitness-map. The website map is interactive so that detailed information for each country is shown with the mouseover function. Not all countries have representative data and therefore caution should be paid when interpreting country comparisons presented this study and in the platform. Data sources are available at: www.fitbackeurope.eu/en-us/fitness-map/sources.

