## Supplementary material for "European Fitness Landscape in Children and Adolescents: updated reference values, fitness maps, and country rankings based on nearly 8 million data points from 34 countries gathered by the FitBack network": Online Supplementary Table 1

**Online Supplementary Table 1.** Summary of the sources used for generating the FitBack reference values and centiles.

|  |  |  |  | Sample size | | | | |  |
| --- | --- | --- | --- | --- | --- | --- | --- | --- | --- |
| **Country** | **Year of testing** | **% male** | **Age (years)** | **BMI** | **WC** | **20mSRT** | **SLJ** | **HGS** | **Reference** |
| Albania | 1994-95 | 53.7% | 10–18 | 2,115 |  |  | 2,114 | 1,984 | Markola et al. 1997 * |
| Austria | 2006-08 | 48.9% | 13–18 | 425 | 415 | 269 | 390 | 388 | [Ortega et al. 2011](https://bjsm.bmj.com/content/45/1/20) |
| Austria | 2015-17 | 48.0% | 8–10 | 204 |  |  | 204 |  | [Greier et al. 2019](https://tp.amegroups.com/article/view/24916/27262) |
| Belgium | 2006-08 | 46.4% | 13–18 | 343 | 337 | 332 | 334 | 337 | [Ortega et al. 2011](https://bjsm.bmj.com/content/45/1/20) |
| Belgium | 2007 | 53.0% | 6–14 | 2,893 | 2,689 | 2,373 | 2,795 | 2,800 | [Vandendriessche et al. 2012](https://journals.humankinetics.com/view/journals/pes/24/1/article-p113.xml) |
| Belgium | 2007-08 | 48.5% | 6–10 |  |  | 650 | 707 | 705 | [De Miguel-Etayo et al. 2014](https://www.nature.com/articles/ijo2014136#Sec2) |
| Belgium | 1997 | 58.0% | 13–18 | 515 |  | 554 | 561 | 262 | [Baquet et al. 2000](https://www.academia.edu/29193085/Effets_dun_cycle_de_course_de_dur%C3%A9e_de_type_intermittent_court_court_sur_la_condition_physique_des_adolescents) * |
| Belgium | 2002? | 48.9% | 10–11 | 591 |  | 591 | 591 | 591 | [Cardon et al. 2004](https://journals.humankinetics.com/view/journals/pes/16/2/article-p147.xml) * |
| Belgium | 1994-04 | 51.9% | 11–18 | 6,493 |  | 6,418 | 6,526 | 6,537 | [Heyters & Marique 2004](https://www.am-sport.cfwb.be/adeps/pdf/Barom%C3%A8tre%20condition%20physique%202011.pdf) * |
| Belgium | 1993-97 | 49.9% | 13–18 | 5,683 |  | 5,535 | 5,660 | 5,690 | [Lefèvre et al. 1998](https://www.lerenbewegenmeten.nl/uploads/eurofit-scores-vlaamse-jeugd.pdf) * |
| Belgium | 2005 | 44.8% | 13–18 | 3,135 |  |  |  |  | [Matton et al. 2007](https://onlinelibrary.wiley.com/doi/10.1002/ajhb.20592) * |
| Belgium | 1994-95 | 52.5% | 13–16 | 2,225 |  | 2,225 | 895 |  | [Telama et al. 2002](https://www.researchgate.net/publication/260096682_Physical_fitness_sporting_lifestyles_and_olympic_ideals_Cross-cultural_studies_on_youth_sport_in_Europe) * |
| Belgium | 2002-04 | 49.1% | 10–13 | 1,055 |  | 1,055 | 1,055 | 1,055 | [Verstraete et al. 2007](https://www.cambridge.org/core/journals/public-health-nutrition/article/comprehensive-physical-activity-promotion-programme-at-elementary-school-the-effects-on-physical-activity-physical-fitness-and-psychosocial-correlates-of-physical-activity/F82D7FD3F51EA985EFBC4116BC4AD3E6) * |
| Bosnia and Herzegovina | 2015-16 | 52.3% | 12–15 | 843 |  | 843 | 843 |  | [Budimlic et al. 2016](https://scindeks-clanci.ceon.rs/data/pdf/0350-3828/2016/0350-38281601055B.pdf) |
| Bulgaria | 1998-99 | 52.5% | 10–18 | 497 |  |  | 497 | 497 | Dimitrova, 2001 * |
| Crete | 2006-08 | 47.4% | 12–18 | 336 | 324 | 192 | 319 | 327 | [Ortega et al. 2011](https://bjsm.bmj.com/content/45/1/20) |
| Croatia | 2015 | 50.5% | 15–18 | 794 | 820 | 596 | 505 |  | [Štefan et al. 2017](https://www.mdpi.com/1660-4601/14/11/1417) |
| Croatia | 2015-18 | 51.6% | 6–8 | 1200 |  |  | 349 |  | [Šalaj et al. 20](http://www.bib.irb.hr/957606/download/957606.iz_zbornika_motoricka_znanja_djece_opis_projekta_norme.pdf)18 |
| Croatia | 2019 | 44.6% | 16–18 | 1,036 | 1,033 |  | 1,036 |  | [Zvonar et al. 2019](https://www.mdpi.com/1660-4601/16/14/2582) |
| Croatia | 2008-09 | 44.8% | 11–18 | 20,616 |  |  | 20,594 |  | [Štefan et al. 2021](https://www.tandfonline.com/doi/abs/10.1080/02701367.2021.1873903) |
| Cyprus | 2007-08 | 51.3% | 6–10 |  |  | 1,111 | 1,173 | 1,184 | [De Miguel-Etayo et al. 2014](https://www.nature.com/articles/ijo2014136) |
| Czechia | 2013-16 | 51.3% | 12–18 | 1,154 |  | 1,086 |  |  | [Rubín et al. 2018](https://doivup.upol.cz/artkey/doi-990003-1500_pohybova_aktivita_a_telesna_zdatnost_ceskych_adolescentu_v_kontextu_zastaveneho_prostredi.php) * |
| Czechia | 1994-95 | 51.0% | 13–16 | 439 |  | 439 | 439 |  | [Telama et al. 2002](https://www.researchgate.net/publication/260096682_Physical_fitness_sporting_lifestyles_and_olympic_ideals_Cross-cultural_studies_on_youth_sport_in_Europe) * |
| Denmark | 2008 | 47.5% | 6–16 | 1,009 | 1,010 |  |  | 5,904 | [Hébert et al. 2020](https://link.springer.com/article/10.1007/s40279-020-01335-3) |
| Denmark | 1996-97 | 39.6% | 16–18 |  |  | 9,342 |  |  | [Nielsen & Andersen, 2003](https://www.sciencedirect.com/science/article/abs/pii/S0091743502000178?via%3Dihub) * |
| Estonia | 2017 | 47.0% | 13–14 |  |  | 142 | 155 | 158 | [Sepp et al. 2017](https://ojs.utlib.ee/index.php/AKUT/article/view/14021) |
| Estonia | 2018- | 49.9% | 8–11 | 212 |  |  | 215 |  | 10.23736/S0022-4707.20.10550-4 |
| Estonia | 2016- | 51.7% | 7–9 | 256 | 256 | 222 | 226 | 226 | [Riso et al. 2019](https://journals.plos.org/plosone/article?id=10.1371/journal.pone.0218901) |
| Estonia | 2016 | 50.9% | 8–9 | 145 | 146 | 136 | 137 | 137 | [Reisberg et al. 2020](https://onlinelibrary.wiley.com/doi/10.1111/sms.13784) |
| Estonia | 2018- | 56.4% | 13–17 | 413 | 413 | 413 | 413 | 413 | [Galan-Lopez et al. 2019](https://www.mdpi.com/1660-4601/16/22/4479) |
| Estonia | 2017 | 51.5% | 12–18 | 3,052 |  | 3,056 | 3,103 |  | <https://www.sportest.eu/> |
| Estonia | 2007-08 | 47.5% | 6–9 |  |  | 725 | 745 | 745 | [De Miguel-Etayo et al. 2014](https://www.nature.com/articles/ijo2014136) |
| Finland | 2013 | 47.6% | 9–15 | 970 | 970 |  |  |  | [Joensuu et al. 2020](https://onlinelibrary.wiley.com/doi/10.1111/sms.13847) |
| Finland | 2007-09 | 51.3% | 9–11 | 374 | 374 |  | 374 |  | [Lintu et al. 2015](https://link.springer.com/article/10.1007/s00421-014-3013-8) |
| Finland | 1995 | 46.6% | 13–16 | 1,109 |  | 1,109 | 1,019 |  | [Telama et al. 2002](https://www.researchgate.net/publication/260096682_Physical_fitness_sporting_lifestyles_and_olympic_ideals_Cross-cultural_studies_on_youth_sport_in_Europe) * |
| France | 2006-08 | 42.2% | 12–17 | 307 | 308 | 258 | 304 | 306 | [Ortega et al. 2011](https://bjsm.bmj.com/content/45/1/20) |
| France | 2009-13 | 49.8% | 9–16 | 9,669 |  | 10,862 |  |  | [Vanhelst et al. 2016](https://www.sciencedirect.com/science/article/abs/pii/S0398762016306162) |
| France | 2010-18 | 51.0% | 6–18 | 31,748 |  |  | 31,748 |  | [Vanhelst et al. 2020](https://onlinelibrary.wiley.com/doi/10.1111/sms.13607) |
| France | 1997 | 50.7% | 12–15 | 507 |  | 507 | 507 | 507 | [Baquet et al. 2001](https://www.thieme-connect.com/products/ejournals/abstract/10.1055/s-2001-14343) * |
| Germany | 2006-08 | 58.9% | 12–18 | 495 | 473 | 392 | 433 | 445 | [Ortega et al. 2011](https://bjsm.bmj.com/content/45/1/20) |
| Germany | 2009-12 | 50.0% | 6–18 | 3,039 | 3,023 |  | 3,043 |  | [Niessner et al. 2020](https://www.frontiersin.org/articles/10.3389/fpubh.2020.00458/full) |
| Germany | 2007-08 | 48.3% | 6–10 |  |  | 638 | 944 | 952 | [De Miguel-Etayo et al. 2014](https://www.nature.com/articles/ijo2014136) |
| Germany | 1994-95 | 51.0% | 13–16 | 977 |  | 977 | 863 |  | [Telama et al. 2002](https://www.researchgate.net/publication/260096682_Physical_fitness_sporting_lifestyles_and_olympic_ideals_Cross-cultural_studies_on_youth_sport_in_Europe) * |
| Greece | 2014 | 51.5% | 6–18 | 306,217 | 304,619 | 176,844 | 256,026 |  | [Tambalis et al. 2015](https://www.tandfonline.com/doi/abs/10.1080/17461391.2015.1088577?journalCode=tejs20) |
| Greece | 2006-08 | 48.6% | 12–18 | 369 | 366 | 346 | 359 | 361 | [Ortega et al. 2011](https://bjsm.bmj.com/content/45/1/20) |
| Hungary | 2006-08 | 49.5% | 13–17 | 397 | 393 | 393 | 394 | 395 | [Ortega et al. 2011](https://bjsm.bmj.com/content/45/1/20) |
| Hungary | 2013 | 51.1% | 10–18 | 580,056 |  | 574,375 | 581,464 | 591,669 | [Csányi et al. 2014](https://www.researchgate.net/publication/264544498_Health-related_Fitness_among_10-18_y_Hungarian_Students_Results_of_a_nationally_representative_study_with_the_Hungarian_National_Student_Fitness_Test_NETFITR?channel=doi&linkId=551c561f0cf20d5fbde5269b&showFulltext=true) |
| Hungary | 2007-08 | 49.6% | 6–10 |  |  | 548 | 1,230 | 1,228 | [De Miguel-Etayo et al. 2014](https://www.nature.com/articles/ijo2014136) |
| Hungary | 1994-95 | 48.7% | 13–16 | 439 |  | 439 | 434 |  | [Telama et al. 2002](https://www.researchgate.net/publication/260096682_Physical_fitness_sporting_lifestyles_and_olympic_ideals_Cross-cultural_studies_on_youth_sport_in_Europe) * |
| Iceland | 2017 | 54.0% | 12–16 | 387 | 387 | 387 | 387 | 387 | [Galan-Lopez et al. 2018](https://www.mdpi.com/1660-4601/15/12/2632/htm) |
| Iceland | 1998 | 51.8% | 10–16 |  |  | 6,130 | 6,202 |  | Gunnarsson & Sigríksson 1999 * |
| Ireland | 2018-19 | 49.9% | 12–16 | 1,147 |  | 1,002 | 1,158 | 1,149 | [O’Keeffe et al. 2020](https://journals.plos.org/plosone/article/peerReview?id=10.1371/journal.pone.0235293) |
| Italy | 2006-08 | 38.8% | 13–18 | 321 | 320 | 263 | 266 | 268 | [Ortega et al. 2011](https://bjsm.bmj.com/content/45/1/20) |
| Italy | 2001-02  2004-05  2007-08  2009-10  2010-11  2011-12 | 55.2% | 6–18 | 4,456 |  |  |  |  | [Lovecchio & Zago, 2019](https://www.minervamedica.it/en/journals/sports-med-physical-fitness/article.php?cod=R40Y2019N02A0298) |
| Italy | 2001-02  2004-05  2007-08  2009-10  2010-11  2011-12 | 48.7% | 12–14 | 6,197 |  |  | 5,898 |  | [Lovecchio & Zago, 2019](https://www.minervamedica.it/en/journals/sports-med-physical-fitness/article.php?cod=R40Y2019N02A0298) |
| Italy | 2001-02  2004-05  2007-08  2009-10  2010-11  2011-12 | 50.9% | 12–14 | 558 |  |  |  |  | [Lovecchio & Zago, 2019](https://www.minervamedica.it/en/journals/sports-med-physical-fitness/article.php?cod=R40Y2019N02A0298) |
| Italy | 2004-13 | 53.0% | 12–16 | 3,331 |  |  | 3,331 |  | Lovecchio et al. 2019 |
| Italy | 2004-13 | 51.5% | 6–18 | 4,376 |  |  | 3,705 |  | Lovecchio et al. 2019 |
| Italy | 2004-13 | 48.4% | 6–18 | 629 |  |  | 510 |  | [Lovecchio et al. 2020](https://www.mdpi.com/1660-4601/17/21/8008/htm) |
| Italy | 2013 | 62.5% | 13–18 | 789 | 722 | 634 | 738 | 770 | [Jemni et al. 2017](https://journals.lww.com/md-journal/Fulltext/2017/12220/Southern_Italian_teenagers__the_older_they_get,.3.aspx) |
| Italy | 2013-14 | 49.5% | 8–10 | 99 |  |  | 99 | 99 | [Colella et al. 2019](https://www.scirp.org/journal/paperinformation.aspx?paperid=92312) |
| Italy | 2007-08 | 50.2% | 6–9 |  |  |  | 1,160 | 1,147 | [De Miguel-Etayo et al. 2014](https://www.nature.com/articles/ijo2014136) |
| Italy | 1997 | 52.9% | 13–18 | 3,638 |  | 3,203 | 3,740 | 3,415 | Cilia et al. 1997 * |
| Kosovo | 2016-17 | 52.8% | 12–18 | 742 |  | 742 | 742 | 742 | [Berisha & Çilli, 2018](https://sportpedagogy.org.ua/index.php/PPS/article/view/853) * |
| Latvia | 2004-09 | 53.6% | 10–18 | 7,743 |  | 3,400 | 7,743 | 7,743 | [Sauka et al. 2010](https://journals.sagepub.com/doi/10.1177/1403494810380298) * |
| Lithuania | 2002, 2012 | 53.6% | 11–18 | 5,339 |  | 5,228 | 5,600 |  | [Venckunas et al. 2018](https://www.frontiersin.org/articles/10.3389/fphys.2018.01797/full) |
| Lithuania | 2016 | 49.8% | 7–11 | 3,214 |  |  | 3,368 |  | [Emeljanovas et al. 2020](https://journals.lww.com/nsca-jscr/Fulltext/2020/02000/Physical_Fitness_and_Anthropometric_Values_Among.16.aspx) |
| Lithuania | 1992 | 46.4% | 12–18 | 3,188 |  | 3,188 | 3,188 | 3,188 | [Jürimäe & Volbekiene, 2006](https://www.tandfonline.com/doi/abs/10.1080/1740898980030206) * |
| Luxembourg | 2003-06 | 55.1% | 9–18 |  |  |  | 1,128 |  | [Woll et al. 2011](https://link.springer.com/article/10.1007/s00431-010-1391-4) |
| Montenegro | 2018-19 | 51.5% | 6–18 | 5,877 | 3,601 |  |  |  | [NCD-RisC, 2020](https://www.sciencedirect.com/science/article/pii/S0140673620318596?via%3Dihub) |
| Netherlands | 2017-19 | 48.9% | 8–14 |  |  |  |  | 1,713 | [Anselma et al., 2021](https://journals.sagepub.com/doi/10.1177/10901981211046533) |
| North Macedonia | 2012 | 51.4% | 6–11 | 1,156 | 1,153 | 1,159 | 1,159 | 1,157 | [Gontarev et al. 2018](https://www.nutricionhospitalaria.org/index.php/articles/01881/show) |
| North Macedonia | 2012 | 51.1% | 10–15 |  |  |  |  | 6,156 | [Gontarev & Ruzdija, 2014](https://www.efsupit.ro/images/stories/nr2.2014/8.%20Art%2028,%20%20pp.%20178%20-%20185.pdf) * |
| Norway | 2004 | 51.9% | 14–16 | 2,604 |  | 2,305 | 2,490 |  | [Haugen et al. 2013](https://journals.sagepub.com/doi/10.1177/1403494813504502) * |
| Poland | 2009-10 | 51.7% | 6–18 | 47,404 |  | 45,925 | 47,326 | 47,061 | [Dobosz et al. 2015](https://cejph.szu.cz/pdfs/cjp/2015/04/11.pdf) |
| Portugal | 2008 | 48.4% | 10–18 | 22,004 | 21,982 | 22,004 |  |  | [Santos et al. 2014](https://www.tandfonline.com/doi/full/10.1080/02640414.2014.906046) |
| Portugal | 2018 | 48.6% | 10–18 | 8,700 | 8,635 | 8,289 | 7,714 | 7,198 | Unpublished data |
| Serbia | 2012-13 | 48.1% | 9–18 | 20,677 |  | 18,778 | 20,341 |  | [Milanovic et al. 2019](https://scielo.isciii.es/pdf/nh/v36n2/1699-5198-nh-36-02-00253.pdf) |
| Slovakia | 1993 | 49.9% | 15–15 | 689 |  | 689 | 689 | 689 | Belej et al.1995 * |
| Slovakia | 1996 | 52.0% | 12–15 | 368 |  | 287 | 323 | 329 | [Kasa & Majherová, 1997](https://www.proquest.com/docview/1306139784?&imgSeq=1) * |
| Slovakia | 1993-95 | 0.0% | 16–16 | 95 |  | 111 | 95 |  | Kyselovicová O. 2000 * |
| Slovakia | 1993 | 59.7% | 10–18 | 3,630 |  | 3,630 | 3,630 | 3,630 | Moravec et al. 1996 * |
| Slovakia | 2014-15 | 51.9% | 10–12 | 426 |  |  | 426 |  | Krska et al. 2015 * |
| Slovenia | 2013-14 | 50.6% | 6–18 | 4,745 | 4,688 | 4,598 | 4,670 | 4,673 | [Morrison et al. 2021](https://www.frontiersin.org/articles/10.3389/fphys.2021.644781/full) |
| Slovenia | 2018 | 50.9% | 6–18 | 210,037 |  |  | 206,804 |  | [Sorić et al. 2020](https://www.nature.com/articles/s41598-020-68102-2#Sec2) |
| Spain | 2006-08 | 48.2% | 12–17 | 413 | 414 | 308 | 397 | 398 | [Ortega et al. 2011](https://bjsm.bmj.com/content/45/1/20) |
| Spain | 2012-20 | 49.5% | 6–18 | 14,645 | 13,952 | 13,450 | 14,129 | 14,155 | [Iglesias-Soler et al. 2021](https://www.frontiersin.org/articles/10.3389/fpsyg.2021.627834/full) |
| Spain | 2018 | 49.4% | 9–11 | 173 | 173 | 171 | 171 | 173 | [Cadenas-Sánchez et al. 2021](https://www.mdpi.com/2072-6643/13/4/1353) |
| Spain | 2017 | 48.1% | 9–12 | 558 | 557 | 551 | 554 | 555 | [Martínez-Vizcaíno et al. 2022](https://onlinelibrary.wiley.com/doi/full/10.1111/sms.14113) |
| Spain | 2013-14 | 46.5% | 6–7 | 518 | 519 | 522 | 519 |  | [Martínez-Vizcaíno et al. 2020](https://bjsm.bmj.com/content/54/5/279) |
| Spain | 2010 | 50.5% | 8–12 | 1,122 |  | 1,061 | 1,116 | 1,118 | [Torrijos-Niño et al. 2014](https://www.jpeds.com/article/S0022-3476%2814%2900181-4/fulltext) |
| Spain | 2019 | 52.0% | 8–16 | 284 | 284 | 284 | 289 | 289 | [Medrano et al. 2020](https://onlinelibrary.wiley.com/doi/10.1111/ijpo.12731) |
| Spain | 2010-11 | 51.5% | 10–18 | 905 | 774 | 889 | 879 |  | Unpublished data |
| Spain | 2011-12 | 51.9% | 6–18 | 2,179 | 2,178 | 2,128 | 2,172 | 2,173 | [Castro-Piñero et al., 2014](https://bmcpublichealth.biomedcentral.com/articles/10.1186/1471-2458-14-400) |
| Spain | 2000-02 | 48.4% | 12–18 | 2,474 | 2,468 | 2,087 | 2,431 | 2,427 | [Ortega et al. 2005](https://www.revespcardiol.org/en-linkresolver-bajo-nivel-forma-fisica-adolescentes-13078126) |
| Spain | 2007-08 | 47.9% | 6–10 |  |  | 38 | 689 | 712 | [De Miguel-Etayo et al. 2014](https://www.nature.com/articles/ijo2014136) |
| Sweden | 2006-08 | 39.8% | 12–18 | 361 | 356 | 255 | 306 | 310 | [Ortega et al. 2011](https://bjsm.bmj.com/content/45/1/20) |
| Sweden | 2007-08 | 48.8% | 6–10 |  |  | 677 | 750 | 659 | [De Miguel-Etayo et al. 2014](https://www.nature.com/articles/ijo2014136) |
| Sweden | 2001 | 51.6% | 11–17 | 1,726 |  |  |  | 1,739 | [Örjan et al. 2005](https://www.tandfonline.com/doi/full/10.1080/11026480500441275) * |
| Switzerland | 2004 | 48.3% | 6–13 | 496 | 491 | 501 |  |  | [Meyer et al. 2014](https://journals.plos.org/plosone/article?id=10.1371/journal.pone.0087929) |
| Switzerland | 1996-97 | 49.5% | 10–18 |  |  | 2,959 | 2,982 |  | Cauderay et al. 2000 * |
| Switzerland | 2005 | 47.9% | 12–12 | 265 |  | 265 |  |  | [Shmid et al. 2007](https://www.yumpu.com/de/document/read/3342862/wie-kann-die-fitness-von-schulkindern-gemessen-werden-sgsm) |
| United Kingdom | 2006-10 | 53.4% | 9–18 | 9,642 | 9,619 | 9,162 |  | 9,397 | [Sandercock et al. 2012](https://www.tandfonline.com/doi/full/10.1080/02640414.2012.660185) |
| United Kingdom | 1999-10 | 51.0% | 10–11 |  |  | 27,954 |  |  | [Boddy et al. 2012](https://journals.lww.com/acsm-msse/Fulltext/2012/03000/Changes_in_Cardiorespiratory_Fitness_in_9__to.16.aspx) * |
| United Kingdom | 2000-03 | 46.5% | 10–13 | 13,152 |  | 3,466 | 13,152 | 13,152 | Ridgers et al. 2006 * |
| United Kingdom | 2009-10 | 51.3% | 10–12 |  |  | 821 | 829 | 824 | [Ranson et al. 2015](https://www.sciencedirect.com/science/article/pii/S0378378215000559?via%3Dihub) * |

20mSRT indicates the 20-m shuttle run test; HGS, handgrip strength; SLJ, standing long jump; BMI, body mass index; WC, waist circumference

*Pseudodata generated from Tomkinson et al. [39].
