## Supplementary material for "European Fitness Landscape in Children and Adolescents: updated reference values, fitness maps, and country rankings based on nearly 8 million data points from 34 countries gathered by the FitBack network": Online Supplementary Table 2

**Online Supplementary Table 2**. Generalized Additive Model for Location, Scale and Shape (GAMLSS) models used to calculate the physical fitness smoothed percentiles.

| **Test** | **Sex** | **Distribution** | **n** | **λ** | **µ** | **σ** | **ν** | **τ** | **SBC** |
| --- | --- | --- | --- | --- | --- | --- | --- | --- | --- |
| 20mSRT | Girls | BCPE | 516811 | 1/3 | 6.16 | 5.30 | 3.94 | 3.39 | 99786154 |
| 20mSRT | Boys | BCPE | 546274 | 1/2 | 6.67 | 5.68 | 4.13 | 3.43 | 123204436 |
| HGS | Girls | BCT | 404897 | 1/2 | 8.26 | 5.01 | 2.94 | 2.66 | 158351826 |
| HGS | Boys | BCT | 422230 | 1/2 | 8.32 | 5.22 | 3.05 | 2.41 | 181302718 |
| SLJ | Girls | BCT | 677639 | 1/2 | 9.60 | 5.42 | 2.84 | 2.19 | 269865621 |
| SLJ | Boys | BCT | 706134 | 1/2 | 9.97 | 5.42 | 2.77 | 2.30 | 286593503 |
| BH | Girls | BCT | 717911 | 1/2 | 15.37 | 5.54 | 2.10 | 2.17 | 211746507 |
| BH | Boys | BCT | 741823 | 1/2 | 15.28 | 5.61 | 2.12 | 2.12 | 229117658 |
| BM | Girls | BCT | 717526 | 1/2 | 9.66 | 5.52 | 2.88 | 2.13 | 225657454 |
| BM | Boys | BCPE | 741678 | 1/2 | 9.61 | 5.61 | 2.95 | 3.73 | 245281509 |
| BMI | Girls | BCT | 716750 | 1/2 | 10.54 | 5.57 | 2.69 | 2.08 | 160587571 |
| BMI | Boys | BCT | 740973 | 1/2 | 10.69 | 5.63 | 2.71 | 2.06 | 170674420 |
| WC | Girls | BCPE | 197832 | 1/2 | 10.62 | 5.20 | 2.38 | 3.37 | 150768194 |
| WC | Boys | BCPE | 205870 | 1/2 | 10.75 | 5.15 | 2.37 | 3.43 | 159088298 |

20mSRT indicates 20-m shuttle run test; HGS, handgrip strength; SLJ, standing long jump; BH, body height; BM, body mass; BMI, body mass index; WC, waist circumference; BCT, Box-Cox t distribution; BCPE, Box-Cox power exponential; SBC, Schwarz Bayesian criterion. Parameters of the fitted distribution are lambda (**λ)**, mu (µ), sigma (σ), nu (ν) and tau (τ)
