## Supplementary material for "European Fitness Landscape in Children and Adolescents: updated reference values, fitness maps, and country rankings based on nearly 8 million data points from 34 countries gathered by the FitBack network": Online Supplementary Table 5

**Online Supplementary Table 5**. Reference values (centiles) for body mass index (kg/m^2^) in European children and adolescents (N=1,464,795)

| Girls  Age (yr.) | P1 | P5 | P10 | P20 | P30 | P40 | P50 | P60 | P70 | P80 | P90 | P95 | P99 |
| --- | --- | --- | --- | --- | --- | --- | --- | --- | --- | --- | --- | --- | --- |
| 6.0-6.9 yrs | 12.3 | 13.2 | 13.7 | 14.3 | 14.9 | 15.4 | 15.8 | 16.4 | 17.0 | 17.8 | 19.1 | 20.4 | 23.4 |
| 7.0-7.9 yrs | 12.4 | 13.3 | 13.8 | 14.6 | 15.2 | 15.7 | 16.3 | 16.8 | 17.5 | 18.5 | 19.9 | 21.4 | 25.1 |
| 8.0-8.9 yrs | 12.6 | 13.6 | 14.2 | 15.0 | 15.7 | 16.3 | 16.9 | 17.5 | 18.3 | 19.4 | 21.0 | 22.7 | 27.0 |
| 9.0-9.9 yrs | 12.9 | 14.0 | 14.6 | 15.5 | 16.2 | 16.8 | 17.5 | 18.2 | 19.1 | 20.2 | 22.0 | 23.9 | 28.7 |
| 10.0-10.9 yrs | 13.1 | 14.2 | 14.9 | 15.9 | 16.6 | 17.3 | 18.0 | 18.8 | 19.7 | 20.9 | 22.8 | 24.8 | 30.0 |
| 11.0-11.9 yrs | 13.4 | 14.6 | 15.4 | 16.3 | 17.1 | 17.8 | 18.5 | 19.3 | 20.3 | 21.5 | 23.6 | 25.7 | 31.3 |
| 12.0-12.9 yrs | 13.9 | 15.2 | 15.9 | 16.9 | 17.7 | 18.5 | 19.2 | 20.0 | 21.0 | 22.3 | 24.4 | 26.6 | 32.3 |
| 13.0-13.9 yrs | 14.6 | 15.9 | 16.7 | 17.7 | 18.5 | 19.2 | 20.0 | 20.8 | 21.8 | 23.1 | 25.2 | 27.4 | 33.2 |
| 14.0-14.9 yrs | 15.2 | 16.5 | 17.3 | 18.3 | 19.1 | 19.8 | 20.6 | 21.4 | 22.4 | 23.6 | 25.7 | 27.9 | 33.8 |
| 15.0-15.9 yrs | 15.5 | 16.9 | 17.7 | 18.7 | 19.5 | 20.2 | 21.0 | 21.8 | 22.7 | 24.0 | 26.0 | 28.2 | 34.1 |
| 16.0-16.9 yrs | 15.8 | 17.2 | 18.0 | 19.0 | 19.8 | 20.5 | 21.2 | 22.0 | 22.9 | 24.2 | 26.2 | 28.4 | 34.5 |
| 17.0-17.9 yrs | 15.9 | 17.3 | 18.1 | 19.1 | 19.9 | 20.6 | 21.3 | 22.1 | 23.0 | 24.3 | 26.3 | 28.5 | 35.0 |
| 18.0-18.9 yrs | 15.9 | 17.4 | 18.1 | 19.1 | 19.9 | 20.6 | 21.3 | 22.1 | 23.0 | 24.2 | 26.3 | 28.6 | 35.4 |
| Boys  Age (yr.) | P1 | P5 | P10 | P20 | P30 | P40 | P50 | P60 | P70 | P80 | P90 | P95 | P99 |
| 6.0-6.9 yrs | 12.7 | 13.5 | 13.9 | 14.5 | 15.0 | 15.5 | 15.9 | 16.4 | 17.0 | 17.8 | 19.1 | 20.4 | 23.7 |
| 7.0-7.9 yrs | 12.7 | 13.6 | 14.1 | 14.7 | 15.3 | 15.8 | 16.3 | 16.8 | 17.5 | 18.4 | 19.9 | 21.4 | 25.4 |
| 8.0-8.9 yrs | 12.9 | 13.8 | 14.4 | 15.1 | 15.7 | 16.3 | 16.9 | 17.5 | 18.2 | 19.2 | 20.9 | 22.7 | 27.4 |
| 9.0-9.9 yrs | 13.2 | 14.2 | 14.8 | 15.6 | 16.2 | 16.8 | 17.5 | 18.2 | 19.0 | 20.1 | 22.0 | 23.9 | 29.3 |
| 10.0-10.9 yrs | 13.4 | 14.4 | 15.1 | 15.9 | 16.6 | 17.3 | 18.0 | 18.7 | 19.6 | 20.8 | 22.9 | 25.0 | 31.1 |
| 11.0-11.9 yrs | 13.6 | 14.7 | 15.4 | 16.3 | 17.1 | 17.8 | 18.5 | 19.3 | 20.3 | 21.6 | 23.8 | 26.1 | 32.7 |
| 12.0-12.9 yrs | 14.0 | 15.1 | 15.9 | 16.8 | 17.6 | 18.3 | 19.1 | 19.9 | 20.9 | 22.3 | 24.5 | 26.9 | 33.8 |
| 13.0-13.9 yrs | 14.4 | 15.7 | 16.4 | 17.4 | 18.2 | 19.0 | 19.7 | 20.6 | 21.6 | 23.0 | 25.3 | 27.7 | 34.5 |
| 14.0-14.9 yrs | 15.0 | 16.3 | 17.0 | 18.1 | 18.9 | 19.6 | 20.4 | 21.3 | 22.3 | 23.6 | 25.9 | 28.3 | 35.1 |
| 15.0-15.9 yrs | 15.5 | 16.9 | 17.7 | 18.7 | 19.5 | 20.3 | 21.1 | 21.9 | 23.0 | 24.3 | 26.6 | 29.0 | 35.7 |
| 16.0-16.9 yrs | 16.0 | 17.4 | 18.2 | 19.3 | 20.1 | 20.9 | 21.7 | 22.5 | 23.5 | 24.9 | 27.2 | 29.5 | 36.2 |
| 17.0-17.9 yrs | 16.3 | 17.7 | 18.6 | 19.6 | 20.5 | 21.3 | 22.1 | 22.9 | 23.9 | 25.3 | 27.5 | 29.9 | 36.6 |
| 18.0-18.9 yrs | 16.5 | 18.0 | 18.9 | 20.0 | 20.8 | 21.6 | 22.4 | 23.2 | 24.2 | 25.6 | 27.8 | 30.2 | 37.0 |

Smoothed percentiles were calculated using the Generalized Additive Model for Location, Scale and Shape (GAMLSS) method and weights were applied according to country population. Age at the midpoint of each interval was selected to provide percentiles. For instance, for the interval 6.0–6.9, data presented were those corresponding to an exact age of a 6.5 year-old child. P10 indicates 10^th^ percentile; other percentiles are abbreviated accordingly. Data sources are available at: [www.fitbackeurope.eu/en-us/fitness-map/sources](http://www.fitbackeurope.eu/en-us/fitness-map/sources)
