## Supplementary material for "European Fitness Landscape in Children and Adolescents: updated reference values, fitness maps, and country rankings based on nearly 8 million data points from 34 countries gathered by the FitBack network": Online Supplementary Table 8

**Online Supplementary Table 8**. Mean percentile and ranking position of each country according to the pooled EU reference values for body mass index and waist circumference.

|  |  | **Body mass index** | | | | |  |  |  |  | **Waist circumference** | | | | |  |
| --- | --- | --- | --- | --- | --- | --- | --- | --- | --- | --- | --- | --- | --- | --- | --- | --- |
|  |  | Both | | Girls | | Boys | |  |  |  | Both | | Girls | | Boys | |
|  | N | Centile | Rank | Centile | Rank | Centile | Rank |  |  | N | Centile | Rank | Centile | Rank | Centile | Rank |
| MCD | 1021 | 58.3 | 1 | 55.1 | 2 | 61.4 | 1 |  | MNE | 3914 | 59.3 | 1 | 59.3 | 1 | 59.3 | 1 |
| SPA | 26024 | 56.3 | 2 | 56.7 | 1 | 56.0 | 2 |  | GRE | 322480 | 55.8 | 2 | 55.6 | 2 | 55.9 | 2 |
| GRE | 324462 | 54.5 | 3 | 53.7 | 5 | 55.2 | 3 |  | ITA | 1061 | 52.6 | 3 | 48.5 | 6 | 55.9 | 3 |
| CRO | 23645 | 53.3 | 4 | 52.6 | 7 | 54.1 | 4 |  | AUS | 416 | 50.6 | 4 | 50.1 | 4 | 51.2 | 4 |
| ISL | 387 | 53.2 | 5 | 54.1 | 3 | 52.3 | 5 |  | POR | 30620 | 50.6 | 5 | 54.1 | 3 | 46.9 | 7 |
| UK | 22794 | 52.1 | 6 | 53.8 | 4 | 50.5 | 10 |  | SPA | 21356 | 49.2 | 6 | 49.1 | 5 | 49.3 | 5 |
| POR | 30707 | 51.8 | 7 | 53.4 | 6 | 50.1 | 13 |  | ISL | 387 | 47.6 | 7 | 47.7 | 8 | 47.6 | 6 |
| HUN | 601480 | 51.6 | 8 | 51.9 | 8 | 51.2 | 7 |  | SWE | 356 | 44.4 | 8 | 47.8 | 7 | 39.1 | 16 |
| SWE | 2087 | 50.4 | 9 | 50.6 | 9 | 50.3 | 12 |  | UK | 9619 | 43.7 | 9 | 44.2 | 9 | 43.3 | 11 |
| IRE | 1147 | 50.2 | 10 | 50.4 | 10 | 49.9 | 15 |  | MCD | 1020 | 42.4 | 10 | 39.1 | 14 | 45.6 | 8 |
| AUS | 630 | 50.0 | 11 | 49.4 | 13 | 50.6 | 9 |  | EST | 815 | 42.2 | 11 | 39.1 | 13 | 44.8 | 9 |
| SVN | 214944 | 49.8 | 12 | 49.6 | 12 | 50.0 | 14 |  | CRO | 1853 | 41.7 | 12 | 39.0 | 15 | 44.7 | 10 |
| SRB | 20677 | 49.7 | 13 | 49.0 | 14 | 50.4 | 11 |  | FIN | 1344 | 40.7 | 13 | 41.4 | 10 | 40.1 | 13 |
| ITA | 26462 | 49.7 | 14 | 48.6 | 16 | 50.7 | 8 |  | GER | 4197 | 40.7 | 14 | 39.5 | 12 | 41.8 | 12 |
| MNE | 6550 | 48.6 | 15 | 45.7 | 21 | 51.5 | 6 |  | SVN | 4848 | 39.6 | 15 | 39.8 | 11 | 39.4 | 15 |
| BEL | 22966 | 48.5 | 16 | 50.1 | 11 | 47.0 | 21 |  | HUN | 393 | 37.9 | 16 | 36.4 | 18 | 39.5 | 14 |
| EST | 4079 | 48.3 | 17 | 47.8 | 18 | 48.8 | 16 |  | FRA | 308 | 36.5 | 17 | 36.4 | 17 | 36.5 | 17 |
| NOR | 2604 | 48.3 | 18 | 48.8 | 15 | 47.9 | 18 |  | DEN | 1042 | 35.2 | 18 | 37.4 | 16 | 32.7 | 19 |
| GER | 5223 | 47.7 | 19 | 47.6 | 20 | 47.8 | 19 |  | BEL | 3059 | 33.7 | 19 | 34.4 | 19 | 33.1 | 18 |
| CZE | 1634 | 47.6 | 20 | 47.8 | 19 | 47.4 | 20 |  | SWI | 492 | 27.6 | 20 | 27.9 | 20 | 27.2 | 20 |
| POL | 49510 | 47.0 | 21 | 45.7 | 23 | 48.3 | 17 |  |  | - | - | - | - | - | - | - |
| BIH | 843 | 45.8 | 22 | 48.0 | 17 | 43.8 | 23 |  |  | - | - | - | - | - | - | - |
| FRA | 42615 | 45.3 | 23 | 45.7 | 22 | 44.8 | 22 |  |  | - | - | - | - | - | - | - |
| FIN | 2453 | 44.0 | 24 | 45.7 | 24 | 42.1 | 28 |  |  | - | - | - | - | - | - | - |
| ALB | 2115 | 43.9 | 25 | 44.8 | 27 | 43.1 | 26 |  |  | - | - | - | - | - | - | - |
| SWI | 762 | 43.7 | 26 | 45.2 | 25 | 42.1 | 29 |  |  | - | - | - | - | - | - | - |
| KOS | 742 | 43.1 | 27 | 45.0 | 26 | 41.4 | 30 |  |  | - | - | - | - | - | - | - |
| LAT | 7743 | 43.1 | 28 | 42.9 | 28 | 43.3 | 25 |  |  | - | - | - | - | - | - | - |
| SLO | 5208 | 41.8 | 29 | 39.6 | 32 | 43.4 | 24 |  |  | - | - | - | - | - | - | - |
| LIT | 11743 | 41.5 | 30 | 40.5 | 30 | 42.4 | 27 |  |  | - | - | - | - | - | - | - |
| DEN | 1041 | 40.7 | 31 | 42.9 | 29 | 38.3 | 32 |  |  | - | - | - | - | - | - | - |
| BUL | 497 | 39.7 | 32 | 40.3 | 31 | 39.1 | 31 |  |  | - | - | - | - | - | - | - |
| N | 1464795 | |  |  |  |  |  |  | N | 409580 | |  |  |  |  |  |

N, sample size and total sample size at the bottom of the table. The 3-digit country codes were used to abbreviate the full country names <https://en.wikipedia.org/wiki/List_of_UNDP_country_codes> as follows: ALB, Albania; AUS, Austria; BEL, Belgium; BIH, Bosnia and Herzegovina; BUL, Bulgaria; CRO, Croatia; CYP, Cyprus; CZE, Czech Republic; DEN, Denmark; EST, Estonia; FIN, Finland; FRA, France; GER, Germany; GRE, Greece; HUN, Hungary; ISL, Iceland; IRE, Ireland; ITA, Italy; KOS, Kosovo; AT, Latvia; LIT, Lithuania; LUX, Luxembourg; NET, Netherlands; MCD, North Macedonia; MNE, Montenegro; NOR, Norway; POL, Poland; POR, Portugal; SRB, Serbia; SLO, Slovakia; SVN, Slovenia; SPA, Spain; SWE, Sweden; SWI, Switzerland; UK, United Kingdom.

For each test, the countries were sorted according to their rank position in the Both (girls and boys) column. Sex- and- age-specific percentile values were calculated using available country-level data and were averaged across sexes and ages to obtain the mean percentile for each country compared to the EU reference values. Smoothed percentiles were calculated using the Generalized Additive Model for Location, Scale and Shape (GAMLSS) method and weights were applied according to country population.
