## Supplementary material for "European Fitness Landscape in Children and Adolescents: updated reference values, fitness maps, and country rankings based on nearly 8 million data points from 34 countries gathered by the FitBack network": Online Supplementary Figure 1

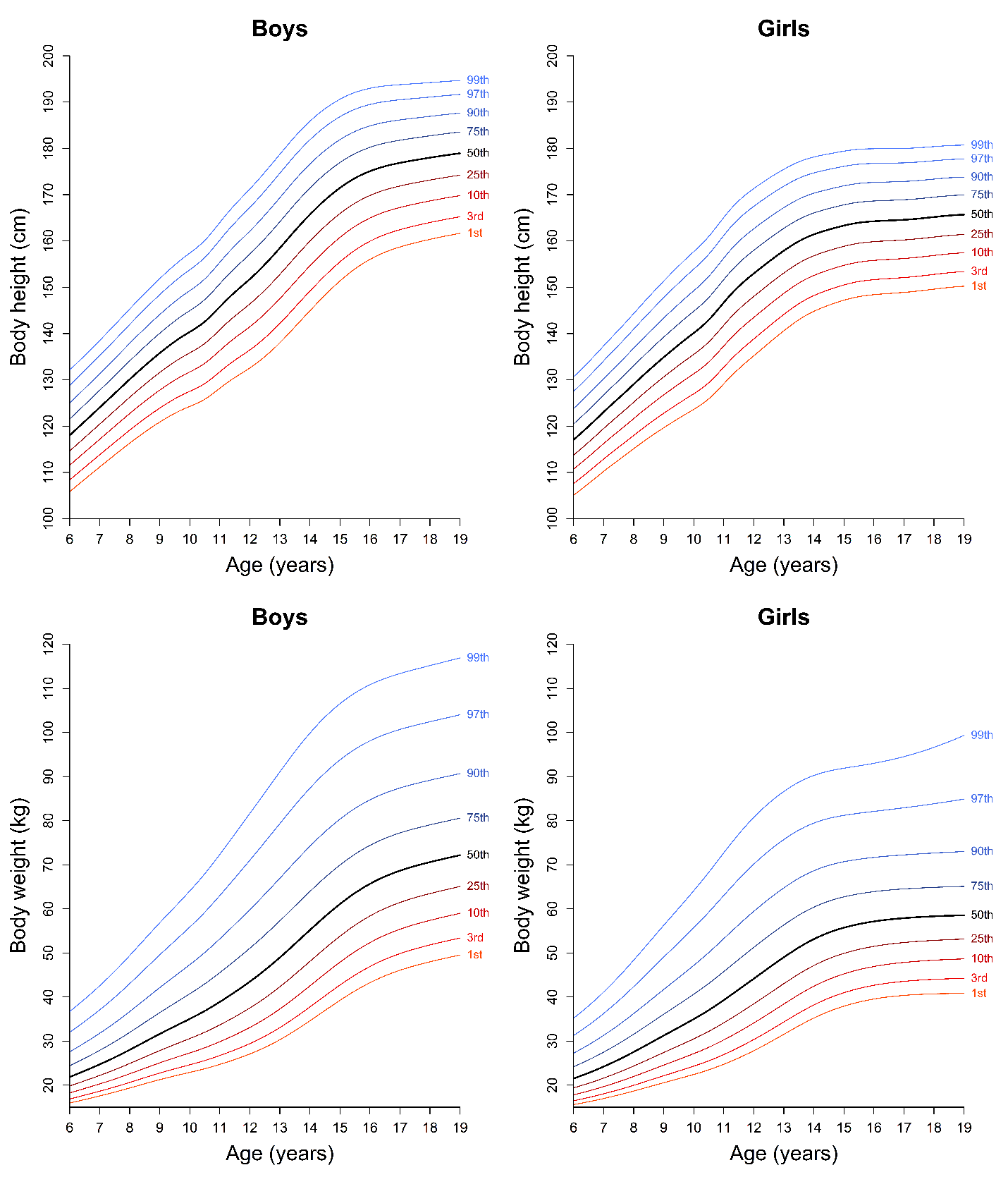

**Online Supplementary Figure 1**. Percentile curves for body height and body mass in European children and adolescents.

Smoothed percentiles were calculated using the Generalized Additive Model for Location, Scale and Shape (GAMLSS) method and weights were applied according to country population. Data sources are available at: <https://www.fitbackeurope.eu/en-us/fitness-map/sources>.
