## Supplementary material for "European Fitness Landscape in Children and Adolescents: updated reference values, fitness maps, and country rankings based on nearly 8 million data points from 34 countries gathered by the FitBack network": Online Supplementary Figure 3

| EU body mass index landscape | EU waist circumference landscape |
| --- | --- |
| 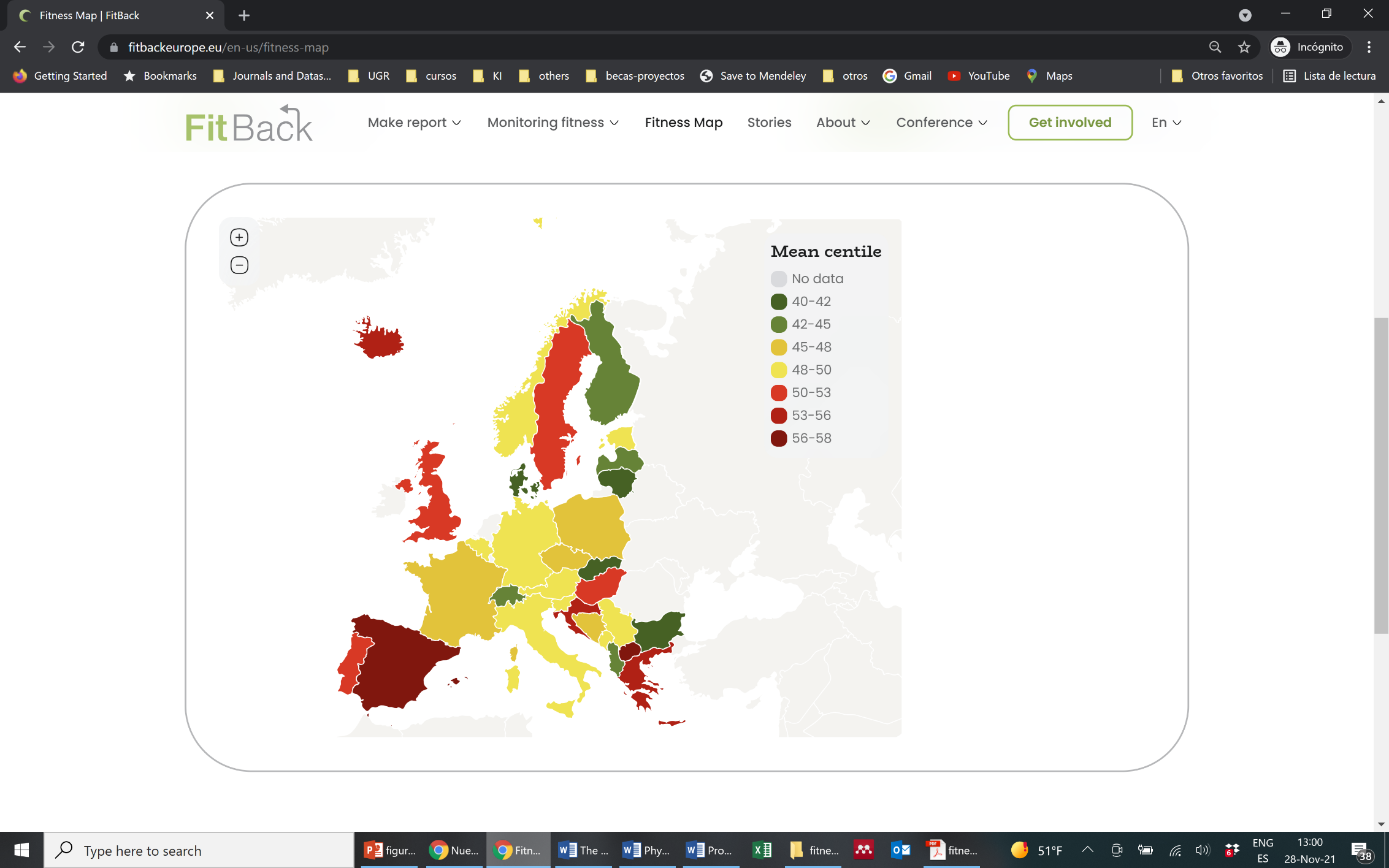 | 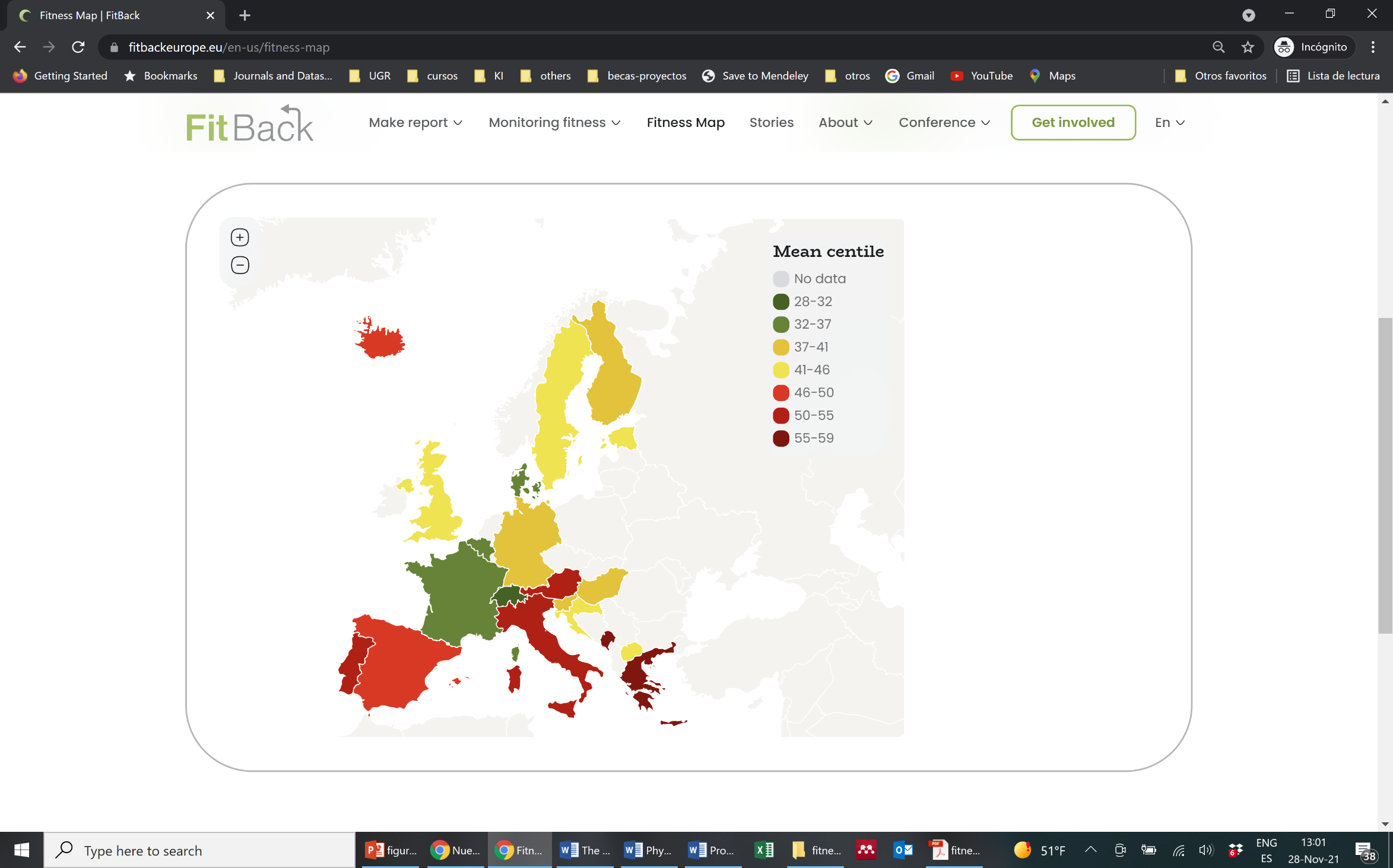 |

**Online Supplementary Figure 3**. European maps for body mass index and waist circumference in children and adolescents.

Sex- and- age-specific percentile values were calculated using available country-level data and were averaged across sexes and ages to obtain the mean percentile for each country compared to the EU reference values. Smoothed percentiles were calculated using the Generalized Additive Model for Location, Scale and Shape (GAMLSS) method and weights were applied according to country population.

Separate European fitness maps for girls and boys are available at: [www.fitbackeurope.eu/en-us/fitness-map](http://www.fitbackeurope.eu/en-us/fitness-map). The website map is interactive so that detailed information for each country is shown with the mouseover function.

Not all countries have representative data and therefore caution should be paid when interpreting country comparisons presented this study and in the platform. Data sources are available at: [www.fitbackeurope.eu/en-us/fitness-map/sources](http://www.fitbackeurope.eu/en-us/fitness-map/sources).
